# Clinical validation of a DNA methylation biomarker to predict overall survival of relapsed ovarian cancer patients

**DOI:** 10.1101/2024.09.18.24312711

**Authors:** Muhammad Habiburrahman, Nahal Masrour, Naina Patel, Anna M Piskorz, Robert Brown, James D Brenton, Iain A McNeish, James M Flanagan

**Author notes:** **Corresponding author** Professor James M Flanagan, Division of Cancer, Department of Surgery and Cancer, Imperial College London. Address : 4th Floor IRDB, Hammersmith Campus of Imperial College London, Du Cane Road, London, United Kingdom, W12 0NN.

## Abstract

**Background:** About 70% of ovarian cancer (OC) patients relapse after initial chemotherapy, making it crucial to predict survival before second-line treatment. Our previous work discovered a blood-based DNA methylation prognostic signature (PLAT-M8) that uses 8 CpG sites related to chemoresistance. We aim to validate this biomarker and its correlation with clinicopathological features and treatment profiles in additional cohorts.

**Methods:** Extracted DNA from whole blood was provided from the BriTROC 1 (n=47) and OV04 cohorts (n=57) upon the first relapse. Additional samples from Hammersmith Hospital (n=100) were collected during first-line chemotherapy (cycles 3-4 and 6). Bisulphite pyrosequencing was used to quantify DNA methylation at the previously identified 8 CpG sites. The methylation data obtained were combined with previous data from ScoTROC 1D and 1V (n=141) and OCTIPS (n=46). Cox regression was used to assess overall survival (OS) after relapse concerning clinicopathological characteristics. The DNA methylation Class (Class 1 vs 2) was determined by consensus clustering.

**Findings:** Blood DNA methylation at relapse predicts better clinical outcomes. Methylation Class shows no association with outcome during first-line chemotherapy treatment. Methylation Class 1 is associated with shorter survival, as indicated by a meta-analysis of five cohorts (OS: HR 2.54, 1.67-3.85). Class 2 patients on carboplatin monotherapy have the best prognosis, while Class 1 patients on the same treatment have the poorest prognosis (OS: aHR 9.69, 2.38-39.47). Class 1 is linked to older patients (>75 years) with advanced-stage, platinum-resistant cases, correlating with residual disease, and shorter progression-free survival. In contrast, Class 2 of PLAT-M8 is linked to platinum-sensitive patients, and higher complete response rates by RECIST criteria, but shows no correlation with CA-125. These findings emphasise the potential of PLAT-M8 in guiding second-line chemotherapy decisions.

**Interpretation:** PLAT-M8 methylation biomarker is associated with survival in OC patients with relapse and hypothetically may predict platinum treatment response at second-line chemotherapy.

**Funding:** This work was supported by funding from Ovarian Cancer Action (“Risk and Prevention” programme grant), Cancer Research UK programme grant (A13086) with support from the Cancer Research UK Imperial Centre, the National Institute for Health Research Imperial Biomedical Research Centre and the Ovarian Cancer Action Research Centre.

**Research in context:** *Evidence before this study:* There is a strong association between platinum-based chemotherapy and DNA methylation changes in blood DNA during ovarian cancer relapse. Previous findings identified eight specific CpG methylation changes (known as PLAT-M8) in blood at relapse following platinum-based chemotherapy that were associated with overall survival in patients enrolled in the ScoTROC 1 trial and the OCTIPS cohort. Using an ovarian cancer cell line model, the study also showed that functional DNA mismatch repair increased the frequency of platinum-induced methylation, providing insights into the observed epigenetic changes.

*Added values of this study:* Our current study validates in five large relapsed ovarian cancer cohorts that: (1) PLAT-M8 is associated with various clinicopathological characteristics, such as age, stage, platinum sensitivity, RECIST response, and progression time; (2) PLAT-M8, particularly from blood samples taken at the time of the first relapse before second-line chemotherapy, can serve not only as prognostic indicators for overall survival but also time to death after relapse in ovarian cancer patients; (3) PLAT-M8 does not have prognostic value when blood samples are taken during first-line chemotherapy before relapse, after initial diagnosis; and (4) PLAT-M8 may stratify overall survival and time to death after relapse based on the second-line treatment received by patients. These findings pave the way for our ongoing research, showcasing the potential of this non-invasive approach in predicting second-line treatment response, guiding decisions, and enhancing outcomes for relapsed ovarian cancer patients.

*Implications of all the available evidence:* The lack of biomarkers guiding treatment decisions during second-line therapy highlights the need for more reliable biomarkers. As a prognostic biomarker, PLAT-M8 is considered simple yet impactful, as it only requires one blood sample taken before second-line treatment at the time of relapse. The advantages of this research include developing personalised treatment approaches, minimizing side effects and wasted time from ineffective medications, reducing the likelihood of subsequent relapse episodes, and improving clinical outcomes for patients. Ultimately, the use of biomarkers has the potential to reduce hospital stays and healthcare costs by optimizing treatment effectiveness and efficiency, while also enhancing the quality of life for patients.

## Introduction

Ovarian cancer (OC) has a high mortality rate in the UK and globally.^1^ Survival rates in the UK are lower than in Asian populations^2^ and other high-income countries, especially Europe.^3^ One reason for the poor survival rate is that over 70% of advanced OC cases relapse after optimal primary surgery and first-line chemotherapy.^4^ With each relapse, the OS decreases, worsening the prognosis.^5^ Platinum-based chemotherapy is a key treatment for both primary and recurrent OC. Currently, the choice of second-line treatment depends on the platinum-free interval (PFI)^6^, which categorises recurrence as sensitive or resistant to platinum-based chemotherapy at the time since the last first-line platinum treatment. This classification guides whether to use platinum monotherapy or in combination with other agents.^6^

A major drawback of classifying platinum sensitivity based on PFI is the risk of biological misinterpretation. This time-based approach ignores the heterogeneous nature of tumorigenesis and does not account for histopathological types like clear cell or mucinous tumours, which are less sensitive to platinum.^7^

Misclassification could affect prognosis and treatment response in relapsed patients receiving second-line platinum. To improve treatment decisions, a more personalised approach using molecular profiling, such as based on epigenetic patterns is needed. Additionally, the current biomarker CA-125 lacks specificity and consistency, underscoring the need for more reliable biomarkers in managing relapsed OC.^8^

Evidence increasingly suggests that epigenetic alterations, such as DNA methylation, contribute to cancer development and can serve as reliable biomarkers for OC.^9,10^ However, the role of epigenetics-based biomarkers in cancer progression and chemotherapy resistance is still lacking sufficient understanding.^9,11^ Epigenetic biomarkers are less invasive than tissue-based ones and can help identify relapse, predict prognosis, and forecast treatment response. However, their clinical use is limited by the complex and time-consuming validation process.^12^ Additionally, a comprehensive analysis of clinicopathological factors is needed to validate prognostic DNA methylation biomarkers in platinum-based treatments. This is crucial to ensure accurate adjustments, as these factors can significantly influence methylation levels and treatment outcomes.^13^ Our previous work has also shown a correlation between a blood-based DNA methylation biomarker (known as PLAT-M8) and OS.^14^ PLAT-M8 is an epigenetic signature constructed by DNA methylation information on eight CpG sites. However, our initial study had a small sample size and did not delve deeper into their associations with clinicopathological characteristics. Thus, further validation is needed. We also aim to investigate its association with survival, second-line treatment stratification, recurrence-related biomarkers, and clinicopathological characteristics in recurrent OC patients in multiple UK cohorts. The ultimate goal is to improve survival rates and reduce the harmful side effects in platinum-nonresponsive patients.

## Methods

### Study design, data source, and samples

This retrospective cohort study aimed to predict OS after cancer relapse by collecting datasets and peripheral blood DNA. Data was obtained from prior cohort studies, including ScoTROC 1V (n=54), ScoTROC 1D (n=87), and OCTIPS (n=46) from Flanagan et al.^14^ who initially used these three datasets to discover PLAT-M8 in 2017. These datasets were originally from ScoTROC 1 (recruited 1998–2000)^15^ and OCTIPS (recruited 1985–2013).^16^ Two additional datasets were added from BriTROC 1 (recruited 2013–2017, n=47)^17^ and OV04 (recruited 2010–2018, n=57).^18^ Furthermore, 153 new blood samples were collected from 100 patients at Hammersmith Hospital (HH) from 2007 to 2014 during first-line chemotherapy cycles (Cycle 3 (n=28), Cycle 4 (n=75), and Cycle 6 (n=49)) to assess the impact of the biomarker before recurrence. The procedures for blood sample collection and patient data collection are outlined in **Figure 1**.

**Figure 1.**
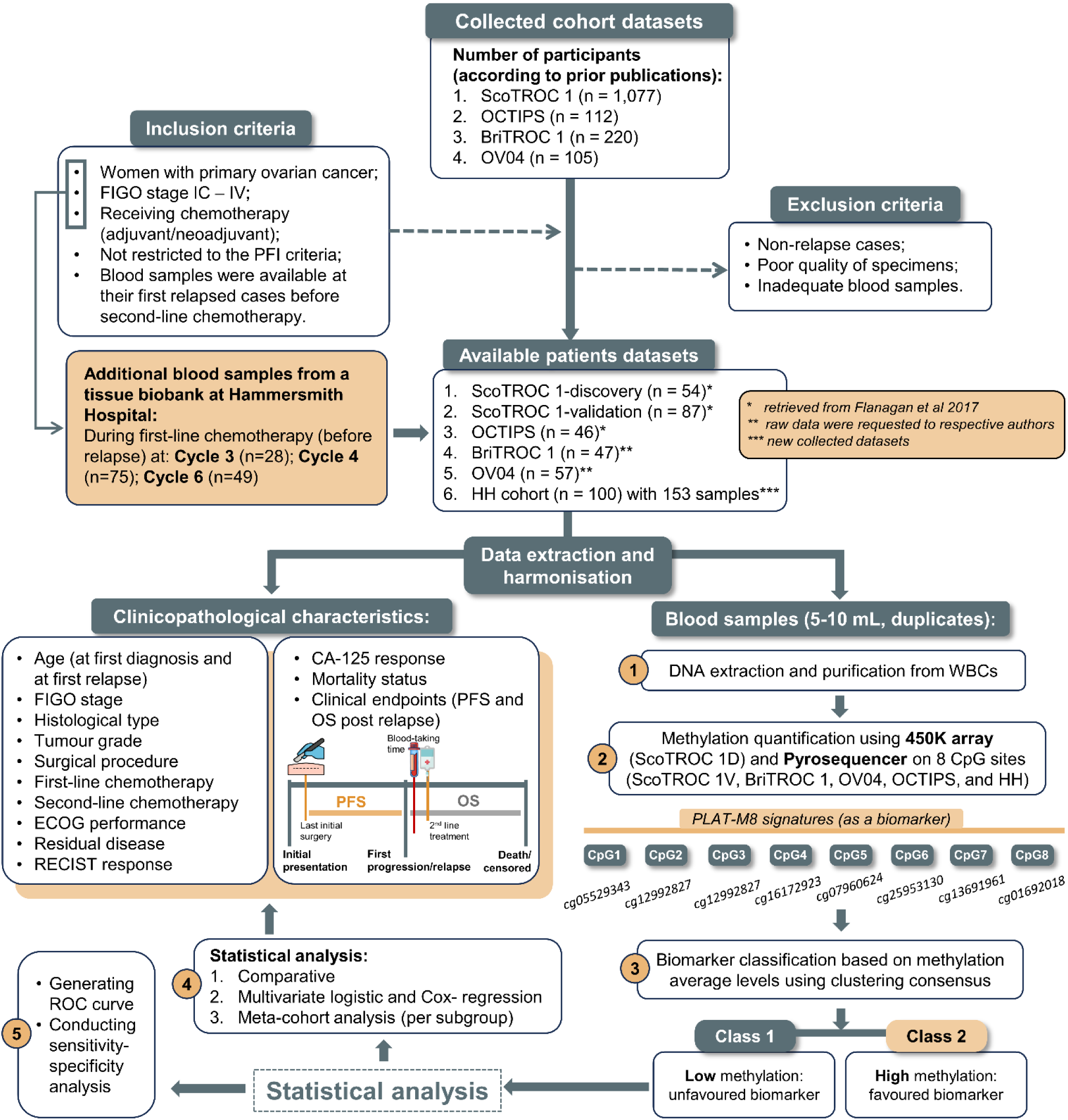
Flow diagram illustrating the data selection process in this study. The study population consists of a subgroup of relapsed ovarian cancer patients registered in 6 datasets of cohorts (ScoTROC-1 is divided into two cohorts, with an additional new collection at Hammersmith Hospital) that were previously studied on a large scale. Inclusion and exclusion criteria were applied to select cases, extract clinical data and analyse stored blood samples for DNA methylation using targeted sequencing with PLAT-M8 epigenetic signatures. PLAT-M8 biomarkers were categorised into two classes based on the level of methylation: class 1 (representing low or absent methylation) and class 2 (representing high methylation levels), which was subsequently analysed for correlation with clinicopathological characteristics. The number of patients (n) at each selection stage and reasons for exclusion in the designated box are provided. ***Abbreviations: BriTROC-1****, British translational research ovarian cancer collaborative 1; **CCC**, Clear cell carcinoma; **CpG**, Cytosine-phosphate-Guanine**; EOC**, epithelial ovarian carcinoma; HGSOC, high-grade serous ovarian carcinoma; **FIGO**, International Federation of Gynecology and Obstetrics; **HH**, Hammersmith Hospital; **OCTIPS**, Ovarian cancer therapy innovative models prolong survival; **OS**, Overall survival after relapse; **OV04**, Ovarian cancer clinical trial study 4^th^ edition; **PFS**, progression-free survival; **PCR**, Polymerase chain reaction; **PFI**, platinum-free interval; **PFS**, Progression-free survival; **ScoTROC-1**, Scottish randomised trial in ovarian cancer (D, discovery and V, validation)*.

### Eligibility criteria

We analysed DNA from blood samples (except for OCTIPS, which was obtained from tumour tissue samples) collected during relapse provided by our collaborators and new DNA extraction from the HH dataset. The present study used specific inclusion criteria to select cases, considering the eligibility criteria established by previous cohorts. These criteria included women diagnosed with primary epithelial OC (including high-grade serous ovarian carcinoma/HGSOC, clear cell carcinoma/CCC, endometrioid carcinoma, and mucinous carcinoma) confirmed at the time of diagnosis, classified as FIGO stage IC– IV, experiencing their first relapse, and receiving platinum. We did not limit the selection based on PFI, and all blood samples were available at the time of their first relapse before second-line chemotherapy. Cases that did not meet the inclusion criteria or had poor sample quality or inadequate blood samples were excluded.

### Clinical variables

This study examined the demographics and clinicopathological profiles of patients with relapsed OC. Recurrence was evaluated using Response Evaluation Criteria in Solid Tumours (RECIST) version 1.1 criteria and CA-125 levels after first-line therapy per Gynaecologic Cancer Intergroup guidelines.^19^ Patient ages at presentation and relapse were categorised per decade, with ’younger patients’ defined as those aged 75 years old or younger, reflecting the peak rate of OC cases in the UK.^20^ Clinical staging was based on FIGO criteria, further divided into early stage (I-II) and advanced stage (III-IV).^21^ Histological types and tumour grades followed World Health Organisation (WHO) criteria regarding serous and non-serous carcinoma.^22^ Chemotherapy regimens were categorised as ’carboplatin monotherapy’ and ’carboplatin with combination’ for first and second-line treatment ^23^. PFI classification was simplified into platinum-sensitive (≥6 months since last chemotherapy) or platinum-resistant (<6 months since last chemotherapy) based on prior research.^24^ Surgical types and outcomes (e.g., residual disease/RD) were simplified, and categorised as either no RD or any RD, accounting for varying cutoffs across all cohorts.^25^ Clinical endpoints included progression-free survival (PFS), and OS after first relapse. The biomarker status of PLAT-M8 was classified into two Classes based on average methylation in 8 CpG sites: Class 1 (hypomethylated or not methylated) and Class 2 (hypermethylated). This classification, determined through clustering consensus as described in a prior publication^14^, was combined with second-line chemotherapy into four categories: Class 2, carboplatin only; Class 2, Other regimens ± carboplatin; Class 1, carboplatin only; and Class 1, Other regimens ± carboplatin.

### Methylation analysis

DNA samples from clinical whole blood were extracted using Qiagen DNA Blood Mini Kits (Qiagen QIAamp®, Manchester, UK) following the manufacturer’s protocols. Quantification was conducted with a Qubit 2.0 fluorimeter (Life Technologies, Paisley, UK) using the dsDNA BR Assay. The methylation status of CpG islands at eight specific sites and these sites was determined in the BriTROC 1, OV04 and Hammersmith Hospital cohorts. Bisulphite conversion of 500 ng of genomic DNA was performed using the EZ-96 DNA-methylation Gold kit (Zymo Research) according to the manufacturer’s instructions. Pyrosequencing was conducted as previously using the primers described.^14^ Consensus clustering identified the optimal two clusters for PLAT-M8 based on cophenetic correlation coefficient assessment.^26^ Samples were classified into Class 1 or Class 2 based on cluster assignment.

### Statistical analysis

The association between clinicopathological factors, mortality status, and biomarker status was assessed using statistical tests such as the χ2, Fisher’s exact, or non-parametric test in SPSS v29. Mann-Whitney and Kruskal-Wallis tests were employed for abnormally distributed data to determine the statistical significance of differences in average age across distinct groups.^27^ Life tables and the log-rank test were used to differentiate the differences between median survival time and rate across cohorts. Furthermore, the factors of age at relapse, FIGO stage, grade, histological type, first-line chemotherapy Class, residual tumour after surgery, and PFS time were selected for analysis in the multivariate logistic regression to identify robustly associated factors linked with Class 1 of PLAT-M8, which is a poorly represented biomarker. The percentages of PLAT-M8 Class 1 and Class 2 were also compared across cohorts using the χ2 test. We aimed to understand which factors contribute to the poorer status of PLAT-M8.^28^

In subsequent statistical analysis, we used R v4.3.1 and RStudio with the following packages: *survminer, survival, survMisc, broom, dplyr, tidyverse, lubridate, ggplot, ggpubr, ggsurvfit,* and *pROC*. The Kaplan-Meier (KM) method was used for survival analysis (OS) in the BriTROC 1 and OV04 datasets, comparing Class-1 and Class-2 groups. The Log-rank test was utilised to determine group differences.^29^ In the ’BriTROC 1 + OV04 study’, OS analysis compared patient groups based on their assigned second-line chemotherapy regimens. Further analysis focused on patients treated with single-agent carboplatin for relapsed disease. No further investigation was done on the ScoTROC 1 and OCTIPS datasets, as research of this nature has already been published elsewhere and no information about second-line chemotherapy.^14^ Biomarker Class 1 of PLAT-M8 versus Class 2 in relapsed and non-relapsed cohorts were separately analysed for hazard ratio (HR) using Cox proportional hazards regression. Biomarker status, along with adjusted clinicopathological factors, were included in a multivariate Cox proportional hazards regression model to predict OS.^28^ At this stage, heterogeneity across different relapsed cohorts was not considered. The HR from univariate Cox proportional hazards regression analysis was assessed across meta-cohorts. Forest plots, employing a random-effects model, were generated to summarise HR for overall effect size estimates in both OS analysis, along with an evaluation of heterogeneity.^30^ Sensitivity analyses of PLAT-M8 in predicting survival after relapse were conducted based on progression time and histological type. In prognostic evaluation, we use time-dependent survival receiver operating characteristic curves (ROCs) to assess cumulative incidence and incidence risk prediction.^31^ Statistical significance was set at p<0.05.

### Ethical approval

Ethical approval for the additional collection of blood samples from patients with OC undergoing first-line chemotherapy from Hammersmith Hospital was granted by the Imperial College Healthcare Tissue Bank (ICHTB) Research Ethics Committee (REC no: 12/WA/0196, project application number: R17016, and ICHTB Human Tissue Authority (HTA) license: 12275) on 2^nd^ May 2017. The study also adheres to ethical standards from prior approved studies, including: (1) ScoTROC 1: Ethical oversight provided by multiple centres, although the specific ethics committee and referral number were not stated^15^; (2) BriTROC 1: Approved by the Cambridge Central Research Ethics Committee, UK (No: 12/EE/0349)^17^; (3) OCTIPS:

Approved by multiple local ethics committees, such as Charité-Universitätsmedizin Berlin (Germany), Medical University of Innsbruck (Austria), Katholieke Universiteit Leuven (Belgium), University of Edinburgh (UK), and other centres that joined this consortium (Nos: EK207/2003, ML2524, 05/Q0406/178, EK130113, 06/S1101/16)^16,32^; (4) OV04: Approved by the National Research Ethics Service (NRES) Committee East of England - Cambridge Central, UK (No: 07/Q0106/63) for Addenbrooke’s Hospital, Cambridge, as well as the Cambridge Central Research Ethics Committee (No: 03/018).^33^ The reporting and writing of this research followed the REporting recommendations for tumour MARKer prognostic studies (REMARK)^34^ and The Strengthening the Reporting of Observational Studies in Epidemiology (STROBE) guidelines.^35^

## Results

### Patient clinicopathological characteristics

We analysed 391 patients and 444 blood samples in this study (**Table S1**). The distribution of PLAT-M8 biomarker Classes varied among cohorts. ScoTROC 1D and HH cycle 6 predominantly had Class 1 biomarker patients, while OCTIPS, BriTROC 1, OV04, and HH samples on cycles 3 & 4 had more Class 2 patients. ScoTROC 1V had a balanced distribution. Patient age and survival time are summarised in **Table 1**, with additional clinicopathological characteristics provided in **Table 2** (mortality status) and **Table S2** (biomarker status). Among 291 relapsed cases (74.42% of the total), age at diagnosis and relapse varied across cohorts. The 2-year rates for relapsed OC patients were 32%. The highest survival rate was in the OCTIPS study, and the lowest was in the ScoTROC 1D study. Non-relapsed patients from the HH study showed moderate survival, with 2-year OS rates of 40%.

**Table 1.** Baseline characteristics of patients in the six included cohorts (n = 391).

| Baseline characteristics | Median (IQR) or median of survival time (95%CI) across different cohorts<sup>a</sup> |  |  |  |  |  |  |  |
| --- | --- | --- | --- | --- | --- | --- | --- | --- |
|  | Relapse |  |  |  |  |  |  | Non-relapse |
|  | ScoTROC 1D | ScoTROC 1V | OCTIPS | BriTROC 1 | OV04 | Overall relapsed cases<sup>b</sup> | p-value<sup>b</sup> | HH |
|  | N = 54 (13.81%) | N = 87 (22.25%) | N = 46 (11.76%) | N = 47 (12.02%) | N = 57 (14.58%) |  |  | N = 100<sup>b</sup> (25.58%) |
| Age at diagnosis (years), n = 391 | 60.50 (52.00-65.00) | 60.00 (53.00-66.00) | 60.00 (53.00-66.00) | 63.00 (53.00-70.00) | 66.00 (55.50-72.00) | 61.00 (53.00-67.00) | **0.004<sup>f</sup>** | 63.50 (54.00-70.00) |
| Age at relapse (years), n = 291 | 61.50 (53.00-65.25) | 61.00 (54.00-67.00) | 61.00 (54.00-67.00) | 65.00 (55.00-73.00) | 69.00 (57.00-74.00) | 62.00 (54.00-69.00) | **0.003<sup>f</sup>** | - |
| Median PFS (months), n = 291 | 10.88 (9.88-11.88) | 12.00 (9.83-14.17) | 28.77 (18.34-39.20) | 24.82 (16.36-33.28) | 22.03 (15.62-28.43) | 16.57 (14.18-18.96) | **<0.001<sup>f</sup>** | - |
| Median OS<sup>c</sup> (months), n = 391 | 9.14 (3.95-14.33) | 13.61 (6.23-20.99) | 50.14 (35.90-64.37) | 18.41 (10.83-25.99) | 21.90 (16.58-27.21) | 19.92 (15.94-23.91) | **<0.001<sup>g</sup>** | 26.10 (18.77-33.44) |
| 2-year OS rate<sup>d</sup>, n = 391 | 18% | 22% | 65% | 35% | 31% | 32% | **<0.001<sup>g</sup>** | 40% |
| Median follow up<sup>e</sup> | 120.33 (37.51-203.15) | 122.47 (50.47-194.46) | 110.20 (79.16-141.24) | 67.86 (64.65-71.07) | 59.87 (39.70-80.03) | 80.71 (62.23-99.19) | **<0.001<sup>g</sup>** | 64.67 (57.80-71.54) |

<sup>a</sup>Data was presented as median (IQR) due to non-normal distribution in some cohorts, while others exhibited normal distribution, prompting homogenization; Survival time: Median (95% CI); and the percentage ‘%’ values represent row percentages.
<sup>b</sup>Only measured the differences of characteristics across the relapsed cases
<sup>c</sup>OS in five cohorts measures the time from the first relapse to death or loss to follow-up (LTFU). For survivors, OS represents the time to their last follow-up (censored time). In the Hammersmith Hospital study (non-relapse cohort), OS specifically tracks the time from the last registered chemotherapy to death or LTFU.
<sup>d</sup>The survival rate was calculated based on life tables, following the cumulative proportion surviving at the end of the interval (5 years).
<sup>e</sup>Median follow-up was calculated using reverse Kaplan Meier from diagnosis before relapse;
<sup>f</sup>Kruskal-Wallis test;
<sup>g</sup>Log rank test
<sup>h</sup>Selected only the first sample from each patient (if there is more than 1 sample given) to avoid duplications in calculating endpoints for the 100 patients as individuals;
**Abbreviations:** *BriTROC 1*, British translational research ovarian cancer collaborative 1; *HH*, Hammersmith Hospital; *OCTIPS*, Ovarian cancer therapy innovative models prolong survival; *OS*, Overall survival after relapse; *OV04*, Ovarian cancer clinical trial study 4<sup>th</sup> edition; *PFS*, progression-free survival; *SCOTROC 1*, Scottish randomised trial in ovarian cancer (D, discovery and V, validation).

**Table 2.** Clinicopathological features in five cohorts with relapsed ovarian cancer cases (n = 291) and at Hammersmith Hospital with non-relapsed cases (n = 100) in relation to mortality status.

| Clinicopathological features | Five cohorts |  |  |  |  |  |  | Hammersmith Hospital cohort |  |  |  |  |  |  |
| --- | --- | --- | --- | --- | --- | --- | --- | --- | --- | --- | --- | --- | --- | --- |
|  | Mortality status |  |  |  | Total |  | p-value | Mortality status |  |  |  | Total |  | p-value |
|  | Alive |  | Death |  |  |  |  | Alive |  | Death |  |  |  |  |
|  | n | % | n | % | n | % |  | n | % | n | % | n | % |  |
| Age at diagnosis (years) |  |  |  |  |  |  |  |  |  |  |  |  |  |  |
| 21-30 | 0 | 0 | 4 | 100 | 4 | 1.4 | 0.112<sup>a</sup> | 0 | 0 | 1 | 100 | 1 | 1.0 | 0.916<sup>a</sup> |
| 31-40 | 1 | 16.7 | 5 | 83.3 | 6 | 2.1 |  | 2 | 40.0 | 3 | 60.0 | 5 | 5.0 |  |
| 41-50 | 13 | 29.5 | 31 | 70.5 | 44 | 15.1 |  | 3 | 23.1 | 10 | 76.9 | 13 | 13.0 |  |
| 51-60 | 22 | 25.3 | 65 | 74.7 | 87 | 29.9 |  | 16 | 66.7 | 8 | 33.3 | 24 | 24.0 |  |
| 60-70 | 28 | 26.9 | 76 | 73.1 | 104 | 35.7 |  | 11 | 32.4 | 23 | 67.6 | 34 | 34.0 |  |
| >70 | 19 | 41.3 | 27 | 58.7 | 46 | 15.8 |  | 10 | 43.5 | 13 | 56.5 | 23 | 23.0 |  |
| **Age at relapse (years)** |  |  |  |  |  |  |  |  |  |  |  |  |  |  |
| 21-30 | 0 | 0 | 3 | 100 | 3 | 1.0 | 0.089<sup>a</sup> |  |  |  |  |  |  |  |
| 31-40 | 1 | 25.0 | 3 | 75.0 | 4 | 1.4 |  |  |  |  |  |  |  |  |
| 41-50 | 10 | 28.6 | 25 | 71.4 | 35 | 12.0 |  |  |  |  |  |  |  |  |
| 51-60 | 23 | 26.1 | 65 | 73.9 | 88 | 30.2 |  |  |  |  |  |  |  |  |
| 60-70 | 24 | 23.3 | 79 | 76.7 | 103 | 35.4 |  |  |  |  |  |  |  |  |
| >70 | 25 | 43.1 | 33 | 56.9 | 58 | 19.9 |  |  |  |  |  |  |  |  |
| **Age at diagnosis category (years)** |  |  |  |  |  |  |  |  |  |  |  |  |  |  |
| Younger ($\leq 75$) | 74 | 27.6 | 194 | 72.4 | 268 | 92.1 | 0.240<sup>b</sup> | 39 | 43.8 | 50 | 56.2 | 89 | 89.0 | 0.350<sup>b</sup> |
| Elder >75) | 9 | 35.3 | 14 | 64.7 | 23 | 7.9 |  | 3 | 27.3 | 8 | 72.7 | 11 | 11.0 |  |
| **Age at relapse category (years)** |  |  |  |  |  |  |  |  |  |  |  |  |  |  |
| Younger ($\leq 75$) | 71 | 27.6 | 186 | 72.4 | 257 | 88.3 | 0.352<sup>b</sup> | | | | | | | |
| Elder >75) | 12 | 39.1 | 22 | 60.9 | 34 | 11.7 |  |  |  |  |  |  |  |  |
| **FIGO stage** |  |  |  |  |  |  |  |  |  |  |  |  |  |  |
| I | 9 | 64.3 | 5 | 35.7 | 14 | 4.8 | **0.023<sup>b</sup>** | 4 | 50.0 | 4 | 50.0 | 8 | 8.0 | 0.716<sup>a</sup> |
| II | 5 | 27.8 | 13 | 72.2 | 18 | 6.2 |  | 6 | 54.5 | 5 | 45.5 | 11 | 11.0 |  |
| III | 55 | 27.5 | 145 | 72.5 | 200 | 68.7 |  | 22 | 37.3 | 37 | 62.7 | 59 | 59.0 |  |
| IV | 14 | 23.7 | 45 | 76.3 | 59 | 20.3 |  | 10 | 45.5 | 12 | 54.5 | 22 | 22.0 |  |
| **FIGO stage degree** |  |  |  |  |  |  |  |  |  |  |  |  |  |  |
| Early (I-II) | 14 | 43.8 | 18 | 56.3 | 32 | 11.0 | **0.043<sup>b</sup>** | 10 | 52.6 | 9 | 47.4 | 19 | 19.0 | 0.297<sup>b</sup> |
| Advanced (III-IV) | 69 | 26.6 | 190 | 73.4 | 259 | 89.0 |  | 32 | 39.5 | 49 | 60.5 | 81 | 81.0 |  |
| **Histological subtypes** |  |  |  |  |  |  |  |  |  |  |  |  |  |  |
| Serous carcinoma | 70 | 34.0 | 136 | 66.0 | 206 | 70.8 | **0.004<sup>a</sup>** | 33 | 42.9 | 44 | 57.1 | 77 | 77.0 | 0.827<sup>a</sup> |
| Adenocarcinoma NOS | 1 | 4.0 | 24 | 96.0 | 25 | 8.6 |  | 0 | 0 | 0 | 0 | 0 | 0 |  |
| Papillary adenocarcinoma | 3 | 14.3 | 18 | 85.7 | 21 | 7.2 |  | 0 | 0 | 0 | 0 | 0 | 0 |  |
| Mucinous adenocarcinoma | 1 | 25.0 | 3 | 75.0 | 4 | 1.4 |  | 0 | 0 | 1 | 100 | 1 | 1.0 |  |
| Endometrioid carcinoma | 6 | 33.3 | 12 | 66.7 | 18 | 6.2 |  | 1 | 33.3 | 2 | 66.7 | 3 | 3.0 |  |
| Clear cell carcinoma | 1 | 20.0 | 4 | 80.0 | 5 | 1.7 |  | 2 | 33.3 | 4 | 66.7 | 6 | 6.0 |  |
| Carcinocarcoma | 0 | 0 | 0 | 0 | 0 | 0 |  | 1 | 50.0 | 1 | 50.0 | 2 | 2.0 |  |
| MCOA histological type | 0 | 0 | 3 | 100 | 3 | 1.0 |  | 3 | 50.0 | 3 | 50.0 | 6 | 6.0 |  |
| Other ovarian malignancies | 1 | 11.1 | 8 | 88.9 | 9 | 3.1 |  | 2 | 40.0 | 3 | 60.0 | 5 | 5.0 |  |
| **Histological group** |  |  |  |  |  |  |  |  |  |  |  |  |  |  |
| Serous carcinoma | 70 | 34.0 | 136 | 66.0 | 206 | 70.8 | **0.001<sup>b</sup>** | 33 | 42.9 | 44 | 57.1 | 77 | 77.0 | 0.751<sup>b</sup> |
| Non-serous carcinoma | 13 | 15.3 | 72 | 84.7 | 85 | 29.2 |  | 9 | 39.1 | 14 | 60.9 | 23 | 23.0 |  |
| **Tumour grade** |  |  |  |  |  |  |  |  |  |  |  |  |  |  |
| Well-differentiated (G1) | 0 | 0 | 0 | 0 | 0 | 0 | 0.760<sup>c</sup> | 2 | 50.0 | 2 | 50.0 | 4 | 4.2 | 0.979<sup>a</sup> |
| Moderately differentiated (G2) | 5 | 45.5 | 6 | 54.5 | 11 | 7.4 |  | 6 | 40.0 | 9 | 60.0 | 15 | 15.6 |  |
| Poorly/undifferentiated (G3-4) | 56 | 40.6 | 82 | 59.4 | 138 | 92.6 |  | 33 | 42.9 | 44 | 57.1 | 77 | 80.2 |  |
| *Missing data* |  |  |  |  | 142 |  |  |  |  |  |  | 4 |  |  |
| **First-line chemotherapy** |  |  |  |  |  |  |  |  |  |  |  |  |  |  |
| Plat monotherapy | 17 | 35.4 | 31 | 64.6 | 48 | 21.0 | 0.174<sup>a</sup> | 9 | 36.0 | 16 | 64.0 | 25 | 25.0 | 0.539<sup>a</sup> |
| Plat + taxane | 36 | 20.5 | 140 | 79.5 | 176 | 76.9 |  | 30 | 44.1 | 38 | 55.9 | 68 | 68.0 |  |
| Plat + TopII inhibitor | 0 | 0 | 0 | 0 | 0 | 0 |  | 1 | 100 | 0 | 0 | 1 | 1.0 |  |
| Plat + ACs | 0 | 0 | 0 | 0 | 0 | 0 |  | 0 | 0 | 1 | 100 | 1 | 1.0 |  |
| Plat + alkylating agents | 1 | 100 | 0 | 0 | 1 | 0.4 |  | 0 | 0 | 0 | 0 | 0 | 0 |  |
| Plat + taxane + STKi | 1 | 50.0 | 1 | 50.0 | 2 | 0.9 |  | 0 | 0 | 0 | 0 | 0 | 0 |  |
| Plat + taxane + EGFR-TKi | 1 | 50.0 | 1 | 50.0 | 2 | 0.9 |  | 0 | 0 | 0 | 0 | 0 | 0 |  |
| Plat + taxane + anti-VEGF | 0 | 0 | 0 | 0 | 0 | 0 |  | 1 | 33.3 | 2 | 66.7 | 3 | 3.0 |  |
| Plat + taxane + TopI inhibitor | 0 | 0 | 0 | 0 | 0 | 0 |  | 0 | 0 | 1 | 100 | 1 | 1.0 |  |
| Plat + taxane + ACs | 0 | 0 | 0 | 0 | 0 | 0 |  | 1 | 100 | 0 | 0 | 1 | 1.0 |  |
| *Missing data* |  |  |  |  | 62 |  |  |  |  |  |  |  |  |  |
| **First-line chemotherapy class** |  |  |  |  |  |  |  |  |  |  |  |  |  |  |
| Cp monotherapy | 17 | 35.4 | 31 | 64.6 | 48 | 21.0 | **0.047<sup>b</sup>** | 9 | 36.0 | 16 | 64.0 | 25 | 25.0 | 0.483<sup>b</sup> |
| Cp with combination | 39 | 21.5 | 142 | 78.5 | 181 | 79.0 |  | 33 | 44.0 | 42 | 56.0 | 75 | 75.0 |  |
| Missing data |  |  |  |  | 62 |  |  |  |  |  |  |  |  |  |
| **Platinum sensitivity level** |  |  |  |  |  |  |  |  |  |  |  |  |  |  |
| Sensitive | 17 | 42.5 | 23 | 57.5 | 40 | 85.1 | **0.039<sup>c</sup>** |  |  |  |  |  |  |  |
| Resistant | 0 | 0 | 7 | 100 | 7 | 14.9 |  |  |  |  |  |  |  |  |
| Missing data |  |  |  |  | 244 |  |  |  |  |  |  |  |  |  |
| **ECOG performance** |  |  |  |  |  |  |  |  |  |  |  |  |  |  |
| 0 | 12 | 22.6 | 41 | 77.4 | 53 | 37.6 | 0.200<sup>b</sup> |  |  |  |  |  |  |  |
| 1 | 8 | 11.1 | 64 | 88.9 | 72 | 51.1 |  |  |  |  |  |  |  |  |
| 2 | 2 | 12.5 | 14 | 87.5 | 16 | 11.3 |  |  |  |  |  |  |  |  |
| Missing data |  |  |  |  | 150 |  |  |  |  |  |  |  |  |  |
| **Surgical type** |  |  |  |  |  |  |  |  |  |  |  |  |  |  |
| Interval debulking | 23 | 44.2 | 29 | 55.8 | 52 | 91.2 | 0.385<sup>c</sup> |  |  |  |  |  |  |  |
| Primary debulking | 1 | 20.0 | 4 | 80.0 | 5 | 8.8 |  |  |  |  |  |  |  |  |
| Missing data |  |  |  |  | 234 |  |  |  |  |  |  |  |  |  |
| **Residual disease** |  |  |  |  |  |  |  |  |  |  |  |  |  |  |
| No residual disease | 30 | 33.0 | 61 | 35.9 | 91 | 39.9 | **0.033<sup>b</sup>** |  |  |  |  |  |  |  |
| Any residual disease | 28 | 20.4 | 109 | 79.6 | 137 | 60.1 |  |  |  |  |  |  |  |  |
| Missing data |  |  |  |  | 63 |  |  |  |  |  |  |  |  |  |
| **RECIST response** |  |  |  |  |  |  |  |  |  |  |  |  |  |  |
| Complete response | 20 | 30.3 | 46 | 69.7 | 66 | 50.0 | 0.149<sup>b</sup> | 27 | 57.4 | 19 | 42.6 | 46 | 47.9 | **0.002<sup>b</sup>** |
| Partial response | 4 | 11.4 | 31 | 88.6 | 35 | 26.5 |  | 5 | 20.8 | 20 | 79.2 | 25 | 26.0 |  |
| Stable response | 4 | 16.7 | 20 | 83.3 | 24 | 18.2 |  | 5 | 31.3 | 11 | 68.8 | 16 | 16.7 |  |
| Progressive disease | 2 | 28.6 | 5 | 71.4 | 7 | 5.3 |  | 1 | 11.1 | 8 | 88.9 | 9 | 9.4 |  |
| Missing data |  |  |  |  | 159 |  |  |  |  |  |  | 4 |  |  |
| **CA-125 response** |  |  |  |  |  |  |  |  |  |  |  |  |  |  |
| Response (decrease) | 13 | 7.4 | 81 | 92.6 | 94 | 77.7 | 0.517<sup>c</sup> |  |  |  |  |  |  |  |
| No response (stable/increase) | 2 | 13.8 | 25 | 86.2 | 27 | 22.3 |  |  |  |  |  |  |  |  |
| Missing data |  |  |  |  | 170 |  |  |  |  |  |  |  |  |  |
| **Second-line chemotherapy** |  |  |  |  |  |  |  |  |  |  |  |  |  |  |
| Plat monotherapy | 11 | 44.0 | 14 | 56.0 | 25 | 17.9 | 0.185<sup>a</sup> |  |  |  |  |  |  |  |
| Taxanes monotherapy | 4 | 66.7 | 2 | 33.3 | 6 | 4.3 |  |  |  |  |  |  |  |  |
| TopI inhibitor monotherapy | 0 | 0 | 4 | 100 | 4 | 2.9 |  |  |  |  |  |  |  |  |
| ACs monotherapy | 2 | 100 | 0 | 0 | 2 | 1.4 |  |  |  |  |  |  |  |  |
| Plat + taxanes | 19 | 38.8 | 30 | 61.2 | 49 | 35.0 |  |  |  |  |  |  |  |  |
| Plat + antimetabolites | 8 | 40.0 | 12 | 60.0 | 20 | 14.3 |  |  |  |  |  |  |  |  |
| Plat + TopI inhibitor | 0 | 0 | 3 | 100 | 3 | 2.1 |  |  |  |  |  |  |  |  |
| Plat + ACs | 9 | 39.1 | 14 | 60.9 | 23 | 16.4 |  |  |  |  |  |  |  |  |
| Taxane + ACs | 0 | 0 | 1 | 100 | 1 | 0.7 |  |  |  |  |  |  |  |  |
| Plat + taxanes + anti-VEGF | 0 | 0 | 3 | 100 | 3 | 2.1 |  |  |  |  |  |  |  |  |
| Plat + taxanes + ACs | 1 | 50.0 | 1 | 50.0 | 2 | 1.4 |  |  |  |  |  |  |  |  |
| Plat + antimetabolites + anti-VEGF | 0 | 0 | 2 | 100 | 2 | 1.4 |  |  |  |  |  |  |  |  |
| Missing data |  |  |  |  | 151 |  |  |  |  |  |  |  |  |  |
| **Second-line chemotherapy class** |  |  |  |  |  |  |  |  |  |  |  |  |  |  |
| Cp monotherapy | 11 | 44.0 | 14 | 56.0 | 25 | 17.9 | 0.538<sup>b</sup> |  |  |  |  |  |  |  |
| Other regimens +/- Cp | 43 | 37.4 | 72 | 62.6 | 115 | 82.1 |  |  |  |  |  |  |  |  |
| Missing data |  |  |  |  | 151 |  |  |  |  |  |  |  |  |  |
| **PFS time for first relapse (days)** |  |  |  |  |  |  |  |  |  |  |  |  |  |  |
| >327 | 68 | 32.9 | 139 | 67.1 | 207 | 71.1 | **0.010<sup>b</sup>** |  |  |  |  |  |  |  |
| ≤327 | 15 | 17.9 | 69 | 82.1 | 84 | 28.9 |  |  |  |  |  |  |  |  |
| **Biomarker status** |  |  |  |  |  |  |  |  |  |  |  |  |  |  |
| Class 2 | 59 | 37.1 | 100 | 62.9 | 159 | 54.6 | **<0.001<sup>b</sup>** | 31 | 46.3 | 36 | 53.7 | 67 | 67.0 | 0.218<sup>a</sup> |
| Class 1 | 24 | 18.2 | 108 | 81.8 | 132 | 45.4 |  | 11 | 33.3 | 22 | 66.7 | 33 | 33.0 |  |

| **Chemo + biomarker status** |  |  |  |  |  |  |  |
| --- | --- | --- | --- | --- | --- | --- | --- |
| Class 2, Cp only | 9 | 69.2 | 4 | 30.8 | 13 | 12.5 | 0.556<sup>a</sup> |
| Class 2, Other regimens +/- Cp | 19 | 38.0 | 31 | 62.0 | 50 | 48.1 |  |
| Class 1, Cp only | 1 | 16.7 | 5 | 83.3 | 6 | 5.8 |  |
| Class 1, Other regimens +/- Cp | 12 | 34.3 | 23 | 65.7 | 35 | 33.7 |  |
| Missing data |  |  |  |  | 187 |  |  |

<sup>a</sup>Mann-Whitney; <sup>b</sup>Chi-Square; <sup>c</sup>Fisher's Exact test.
The percentage '%' values in each group represent row percentages; meanwhile, the percentage '%' values in the total represent column percentages.
**Abbreviation:** **ACs**, anthracyclines; **Cp**, carboplatin; **ECOG**, Eastern cooperative oncology group; **EGFR-TKi**, epidermal growth factor receptor tyrosine kinase inhibitor; **FIGO**, International federation of gynecology and obstetrics; **MCOA**, Mixed cell ovarian adenocarcinoma; **NOS**, non-specific; **Plat**, platinum; **RECIST**, Response evaluation criteria in solid tumors; **STKi**, serine-threonine kinase inhibitors; **TopII**, topoisomerase II; **VEGF**, Vascular endothelial growth factor. **Notes:** +/- Cp' means with or without carboplatin since some patients did not receive Carboplatin as their primary therapy, and 'other' means other regimens of chemotherapy beside carboplatin. Taxane (e.g., paclitaxel and docetaxel), TopII inhibitor (e.g., etoposide), ACs (e.g., liposomal doxorubicin and epirubicin), Alkylating agents (e.g., cyclophosphamide), STKi (e.g., enzastaurin), EGFR-TKi (e.g., erlotinib and sorafenib), anti-VEGF (e.g., bevacizumab), TopI inhibitor (e.g., topotecan), Antimetabolites (e.g., gemcitabine)

Across five relapsed-cases cohorts, significant differences were observed in PFS, OS, and follow-up months. Data from non-relapsed patients at HH (n=100, 25.58% of total) showed a slightly older median age at diagnosis, with longer OS, but shorter follow-up months compared to relapsed patients. Our relapsed patients were relatively more dominant in later stages, with 89.0% in stages III-IV and 11.0% in stages I-II. The main histological type was serous carcinoma (70.8%), followed by adenocarcinoma NOS (8.6%) and papillary adenocarcinoma (7.2%). Most patients (92.6%) had poorly differentiated and undifferentiated (G3-G4) tumours.

For first-line chemotherapy, 76.9% of patients received predominantly platinum and taxane (carboplatin + paclitaxel). Other regimens with or without carboplatin constituted the greater proportion of 229 available patient data from ScoTROC 1D and 1V, OCTIPS, and BriTROC 1. In second-line treatment, 35.0% of patients predominantly received platinum and taxane (carboplatin + paclitaxel), and other regimens with or without carboplatin constituted the majority (72.9%) in 140 available data from OCTIPS, BriTROC 1, and OV04. In ScoTROC 1D and 1V, 51.1% of patients were in Eastern Cooperative Oncology Group (ECOG) grade 1, meaning they had restricted physically strenuous activity but were ambulatory and capable of light or sedentary work. Most patients (91.2%) underwent interval debulking surgery and about 60.1% of patients had RDs.

However, 50% of patients achieved a complete response per RECIST, and 77.7% had a good response (decreasing CA-125 levels). PLAT-M8 biomarker Classes were relatively balanced in the relapsed cohort (Class 2: Class 1 ratio = 6:5) but slightly favoured Class 2 in the non-relapsed cohort (ratio = 7:5). Specifically, in blood biomarker datasets, 48.1% in Class 2 and 33.7% in Class 1 received other regimens with or without carboplatin as their second-line chemotherapy.

### Association between clinicopathological features with mortality and biomarker status

In a comparative analysis of mortality status among relapsed cases (**Table 2**), significant differences were observed in various clinicopathological features: (1) Advanced FIGO stage correlated with a higher death rate compared to earlier stages (p=0.043); (2) Non-serous carcinoma patients had a higher death rate than serous carcinoma patients (p=0.001); (3) Patients who received other regimens with or without carboplatin in first-line chemotherapy had a higher death rate than those with carboplatin only (p=0.047); (4) Platinum-resistant patients had a higher death rate than platinum-sensitive individuals (p=0.039); (5) Patients with residual disease (RD) had a higher death rate than those without RD (p=0.033); (6) Shorter PFS time in relapsed individuals correlated with higher mortality compared to longer PFS time (p=0.010); and (7) Patients with biomarker Class status Class 1 of PLAT-M8 at relapse had more deaths than those in Class 2 (p<0.001). Conversely, in a comparative analysis of mortality status among non-relapsed cases in the HH cohort, no significant differences were found in clinicopathological features. However, there was a notable distinction in RECIST response, with a higher percentage of deceased patients showing evidence of progressive disease (p=0.002).

In analysing biomarker status among relapsed cases (**Table S2**), significant variations in clinicopathological features were noted: (1) Patients over 75 had a higher percentage in Class 1 than younger patients (p=0.015); (2) Advanced FIGO stage patients had a higher proportion in Class 1 than earlier stages (p=0.038); (3) Patients with RD had a higher percentage of Class 1 biomarker than those without RD (p=0.024); (4) the shorter PFS time in relapsed individuals correlated with Class 1 of PLAT-M8 (p<0.001), as well as PLAT-M8 association with platinum sensitivity and RECIST. Conversely, the comparative analysis based on biomarker status among non-relapsed cases in the HH cohort showed no significant differences in clinicopathological features. In exploring the association between the methylation-based biomarker and clinicopathological features, this study employed multivariate logistic regression (**Table S3**). It particularly assessed the prediction of Class 1 PLAT-M8 biomarkers, known for unfavourable clinical outcomes. Before adjustment, the FIGO stage, RD, and PFS contributed to PLAT-M8 status. Post-adjustment, only PFS significantly predicted Class 1, with an odds ratio of 6.33 (3.12-12.84), p<0.001.

### PLAT-M8 validation in BriTROC 1 and OV04 patients

Our previous research on DNA methylation in blood, utilising ScoTROC 1 datasets for training and validation, was further validated against OCTIPS tissue biopsies. The proposed PLAT-M8 biomarker, an epigenetic signature from 333 CpG sites, is associated with survival outcomes with a false discovery rate (FDR) of less than 10%.^14^ After incorporating BriTROC 1 and OV04 cohorts, data normalisation accommodated methylation analysis variations (Pyrosequencing and 450k methylation array). **Figure S1** illustrates methylation analysis in blood samples from relapsed OC patients across four cohorts. Remarkably, ScoTROC 1D-450K displayed decreased methylation levels after normalisation, aligning closely with the other three cohorts. In this study, we clinically validated PLAT-M8 in BriTROC 1 and OV04 cohorts. Adjusted Cox regression model from BriTROC 1 revealed poorer OS for methylation Class 1 compared to Class 2 with aHR of 2.92 (1.21-7.02, p=0.017) (**Figure 2A**). OV04 datasets showed no difference in OS between Class 1 and Class 2 (**Figure 2B**). When the two datasets were combined, similar patterns persisted, with Class 1 exhibiting poorer OS (aHR 2.21, 1.24-3.91, p=0.007) (**Figure 2C**). Examining methylation changes between PLAT-M8 biomarker Class 1 and Class 2 using a pyrosequencer specific to CpG sites (**Figure S2**), in BriTROC 1, PLAT-M8 showed significant hypermethylation at 4 out of 8 CpG sites and hypomethylation at 1 out of 8 CpG sites. Similar patterns were observed in OV04, with 4 out of 8 CpG sites hypermethylated and 1 out of 8 CpG sites hypomethylated.

**Figure 2.**
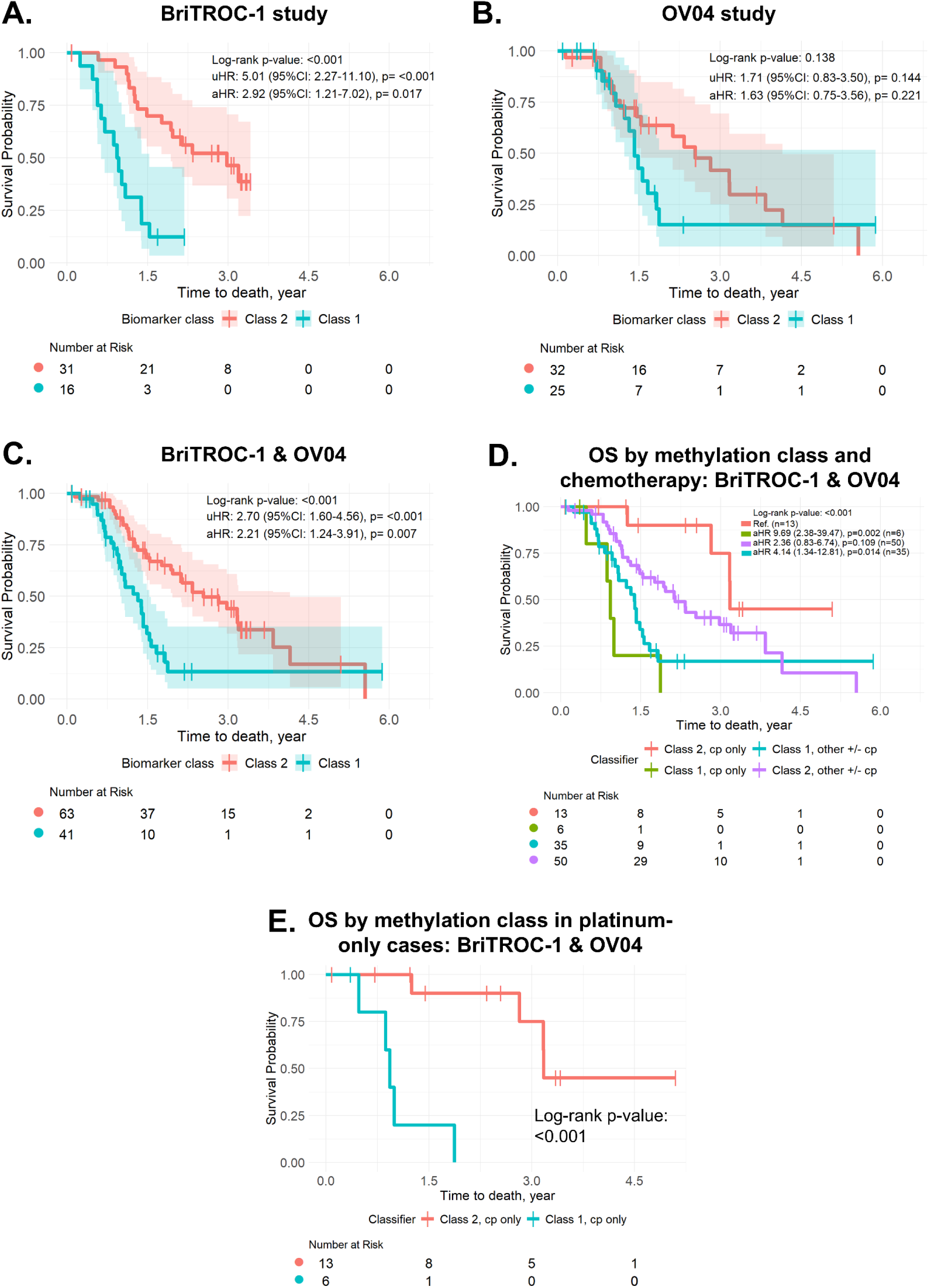
PLAT-M8 validation in BriTROC-1 and OV04 patients reveals prognostic methylation markers for overall survival (OS) after first relapse and methylation markers at relapse potentially predict second-line platinum-based chemotherapy response. **(A)** In BriTROC-1 (n = 16, Class 1 in blue; n = 31, Class 2 in red), Kaplan-Meier (KM) curve shows that Class 1 biomarker status has a poorer prognosis than Class 2 (reference) with adjusted multivariable Cox regression for OS at adjusted hazard ratio (aHR) 2.92 (95%CI: 1.21-7.02, p = 0.017), log-rank p < 0.001. **(B)** In OV04 (n = 25, Class 1 in blue; n = 32, Class 2 in red), KM curve indicates a nonsignificant tendency toward a poorer prognosis for relapse methylation between Class 1 and Class 2. Adjusted multivariable Cox regression for OS at aHR 1.63 (95%CI: 0.75-3.56, p = 0.221), log-rank p = 0.138. **(C)** Adjusted multivariable Cox regression for OS is aHR 2.21 (95%CI: 1.24-3.91, p = 0.007), log-rank p < 0.001. All adjustments were made for the covariates of age at relapse, cancer stage, histology, and progression-free survival (PFS). **(D)** Data from BRITROC-1 and OV04 were combined, and a KM curve for OS was generated for each class, stratified by second-line treatment. The survival curves analysed the clinical outcomes of patients who received carboplatin (Cp) only, either as a single agent in the Class 1 biomarker group (“Class 1, Cp only”) or the Class 2 biomarker group (“Class 2, Cp only”). Other treatments, with or without Cp, were also analysed within Class 1 (“Class 1, other +/- Cp,” n = 35, green) and Class 2 (“Class 2, other +/- Cp,” n = 50, purple), including paclitaxel (n = 56), liposomal doxorubicin (n = 21), gemcitabine (n = 9), cediranib (n = 3), epirubicin (n = 2), and bevacizumab (n = 2). In this analysis, “Class 2, Cp only” was used as the reference group. The OS post-relapse analysis indicated that “Class 2, Cp only” had the most favourable prognosis, while “Class 1, Cp only” had the poorest prognosis, with an aHR of 9.69 (95% CI: 2.38-39.47, p = 0.002). The “Class 1, other +/- Cp” and “Class 2, other +/- Cp” groups showed intermediate results (overall log-rank p < 0.001). **(E)** Comparing the OS between those who received Cp only in Class 1 (n = 6) versus Class 2 (n = 13), the “Class 2, Cp only” group showed a more favourable prognosis (log-rank p < 0.001). All charts were truncated at 6 years for consistency across studies.

### PLAT-M8 detection at relapse as a predictor of survival: Validation in large cohorts

Validating in five datasets, multivariate Cox regression (**Table 3**, **Figure 3A**) revealed that Class 1 of PLAT-M8 predicted poorer OS compared to Class 2 (aHR 1.82, 1.35-2.46, p<0.001). Other prognostic factors for OS included FIGO advanced stage (aHR 1.87, 1.13-3.08, p=0.014), non-serous histological type (aHR 1.82, 1.33-2.50, p<0.001), and PFS ≤327 days (aHR 1.87, 1.33-2.64, p<0.001).

**Figure 3.**
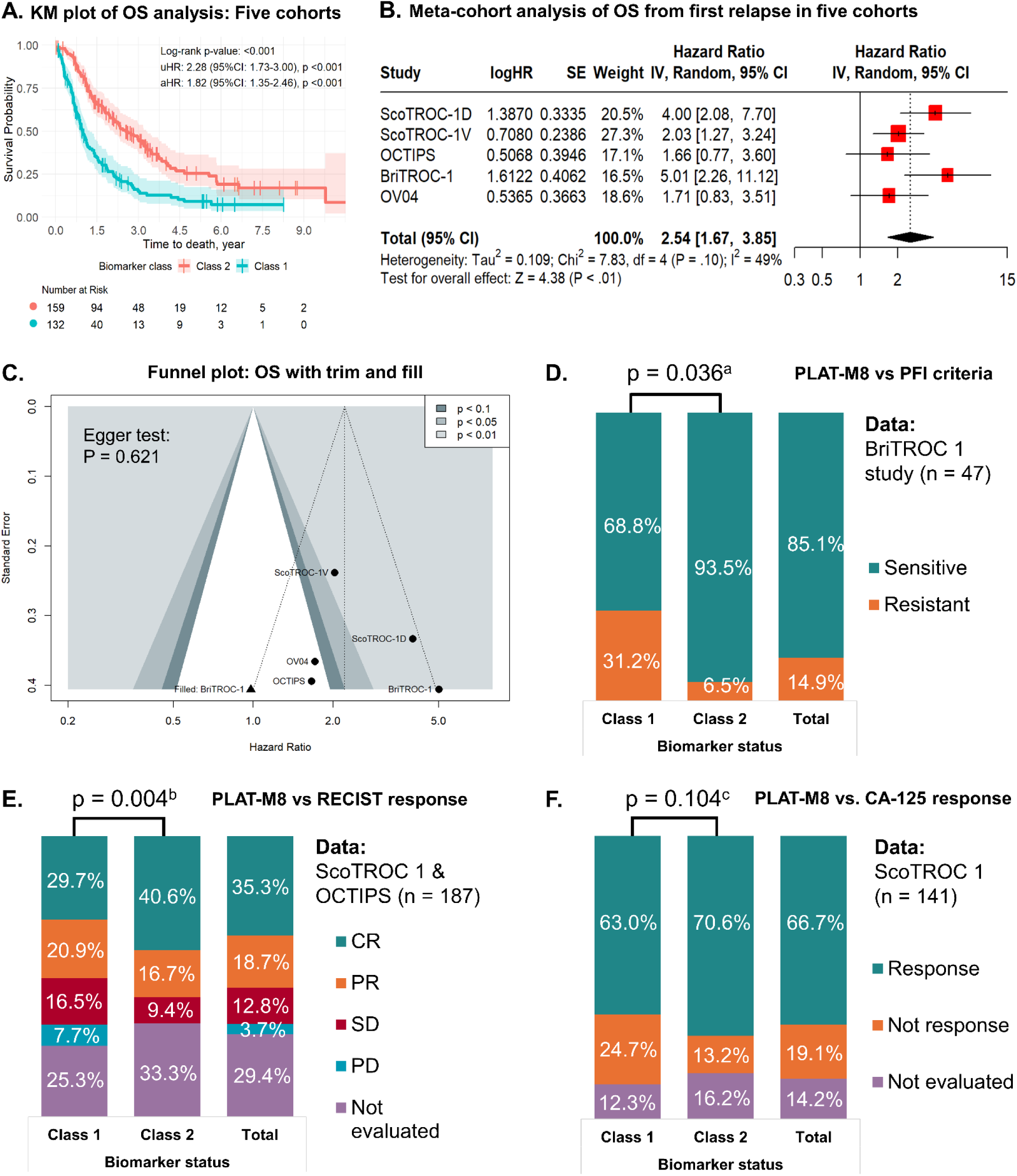
PLAT-M8 validation in five cohorts (ScoTROC-1D, ScoTROC-1V, and OCTIPS from Flanagan et al. 2017, and addition of BriTROC-1 and OV04 studies) along with newly collected samples from Hammersmith Hospital (HH) with cycle-specific data. In 291 patients across the five cohorts, Kaplan-Meier curves for relapse methylation Class 1 (blue, n = 138) versus Class 2 (red, n = 153) revealed **(A)** multivariable adjusted Cox regression for overall survival (OS) adjusted hazard ratio (aHR) of 1.82 (95%CI: 1.35-2.46, p < 0.001), log-rank p < 0.001. All adjustments were made for the covariates of age at relapse, cancer stage, histology, and progression-free survival (PFS). Considering heterogeneity, meta-cohorts with a forest plot of univariate Cox regression showed **(B)** Meta-cohorts with a forest plot of univariate Cox regression using a random-effects model for OS analysis among relapse cases showed that Class 1 of PLAT-M8 had poorer prognosis with a summary HR of 2.54 (95%CI: 1.67-3.85, p < 0.01) with possible moderate heterogeneity (I^2^=49%, p = 0.10). **(C)** Contoured funnel plot evaluation of publication bias in meta-cohorts of 5 datasets of relapsed cases for OS analysis. The contour-enhanced funnel plot incorporates three shaded areas of statistical significance. The majority of the experiments fall within the light grey area (highly significant results, p < 0.01). According to the Egger’s test, there was no significant publication bias, and the plot exhibits relative symmetry despite the small number of studies. Visual inspection of the plot symmetry using the trim and fill analysis would impute potential missing studies close to the threshold of significance within the grey area of statistical significance. The trim and fill analysis revealed a study (indicated by a black triangle) in the OS meta-cohort. Additionally, PLAT-M8 was validated with recurrence-related parameters such as platinum sensitivity (PFI), RECIST response, and CA-125 response. **(D)** Histogram from BriTROC-1 samples, which were only available for assessing the PFI (Class 1, n = 16 and Class 2, n = 31), showed that Class 2 associated with platinum sensitivity (^a^Fisher’s Exact test, p = 0.036). **(E)** Histogram from ScoTROC-1 and OCTIPS samples, which were only available for assessing the RECIST response (Class 1, n = 68 and Class 2, n = 64), revealed that Class 2 had a higher proportion of patients with complete response (^b^Mann-Whitney test, p = 0.004). **(F)** Histogram from ScoTROC-1 samples, which were only available for assessing the CA-125 response (Class 1, n = 64 and Class 2, n = 57), indicated a tendency for Class 2 to have a higher proportion of responsive patients (^c^ χ2 test, p = 0.104).

**Table 3.**
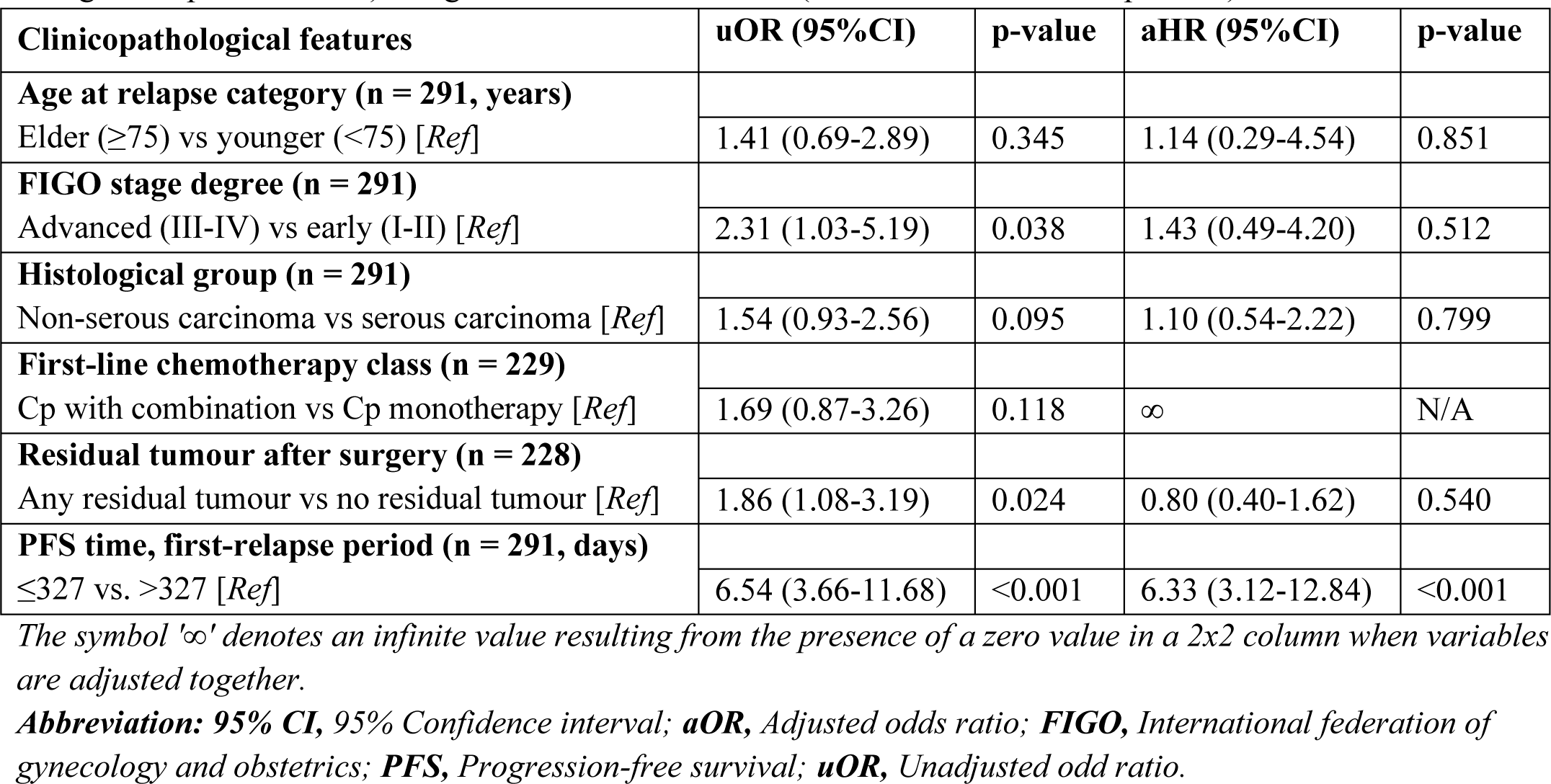
Multivariate Cox regression analysis associating overall survival after relapse with biomarker class, adjusted by clinical covariates in five cohort datasets (n = 291; only complete data variables).

After performing univariate Cox regression in each cohort, we combined all relapsed-cases cohorts across studies using both blood and tissue biopsy samples. A meta-cohort analysis was conducted using a random-effect model, accounting for heterogeneity in OS (**Figure 3B**). This analysis yielded a higher summary HR of 2.54 (1.67-3.85, p<0.01) compared to the previous combined analysis without meta-analysis.

Differences in survival between PLAT-M8 Classes, especially within the first 2 years, were observed for OS in relapse cases (**Table S4**). Notable variations in median PFS and OS were identified across diverse clinicopathological characteristics, revealing reduced median survival (p<0.001 for all three parameters) in Class 1 of PLAT-M8 during relapse compared to Class 2 (**Table S5**). Blood DNA methylation at relapse predicted clinical outcomes, with the better prognosis group (Class 2 of PLAT-M8) showing a median difference of 18.8 months in OS after relapse and more than 1.6 times that of the poorer group. Furthermore, advanced stage (III-IV) was associated with significantly diminished median PFS (p=0.004) and OS (p=0.026), compared to earlier stage (I-II), along with abbreviated median survival in non-serous carcinoma relative to serous carcinoma (p<0.001 for all three parameters). Additional findings included decreased median PFS (p=0.003) in patients receiving regimens excluding carboplatin during first-line treatment, reduced median survival in platinum-resistant patients versus sensitive populations (p<0.001 for all three parameters), and diminished median survival in patients with any residual diseases compared to those without (p<0.001 for all three parameters). Additionally, patients with no response to CA-125 experienced significantly reduced median PFS (p=0.004) compared to those with a positive response.

Furthermore, we retrospectively explored PLAT-M8’s association with recurrence-related parameters (platinum sensitivity, RECIST response, CA-125 response). PLAT-M8 methylation showed a significant link with platinum sensitivity after first-line chemotherapy, indicating Class 1 as more resistant and Class 2 as more sensitive (p=0.036, **Figure 3D**). Class 1 predominated in PD (100%), SR (62.5%), and PR (54.3%) compared to CR (40.9%) with p=0.004 (**Figure 3E**). Although not significantly associated, there was a tendency for more responsive patients to be in Class 2 for CA-125 response (p=0.104, **Figure 3F**). **Figure 3C** assessed publication bias in OS using a funnel plot. Despite a small number of studies, relative symmetry was observed, and Egger’s test found no significant bias (p>0.05). Trim and fill analysis suggested potential missing studies in OS, influencing overall results. It is worth noting that some prior studies (BriTROC 1 and OV04) did not originally focus on PLAT-M8, and the HH cohort provided new datasets for this validation study.

### PLAT-M8 prognostic role based on the treatment regiments

Patients were categorised based on their second-line treatment regimens in the BriTROC 1 and OV04 cohorts (**Table S6**). Notably, Class 2 patients receiving carboplatin monotherapy had the best prognosis in OS, while Class 1 patients receiving carboplatin monotherapy had the poorest prognosis (aHR 9.69, 2.38-39.47, p=0.002) (**Figure 2D**)). Further analysis of patients who received platinum monotherapy (carboplatin only) in **Figure 2E** revealed a more favourable prognosis for patients in Class 2 who received carboplatin monotherapy in OS (log-rank p<0.001).

### PLAT-M8 lacks predictive value during initial chemotherapy cycles

Additional data from 100 OC patients in the HH cohort undergoing chemotherapy was collected to investigate the role of PLAT-M8 before relapse. Comparative analysis (**Table 2, Table S2**) showed no significant differences in clinicopathological features related to mortality and biomarker status. Multivariate Cox regression (**Table 4**, **Figure S3**) also did not reveal significant predictive roles for PLAT-M8 in OS, considering age, FIGO stage, and histological group.

**Table 4.**
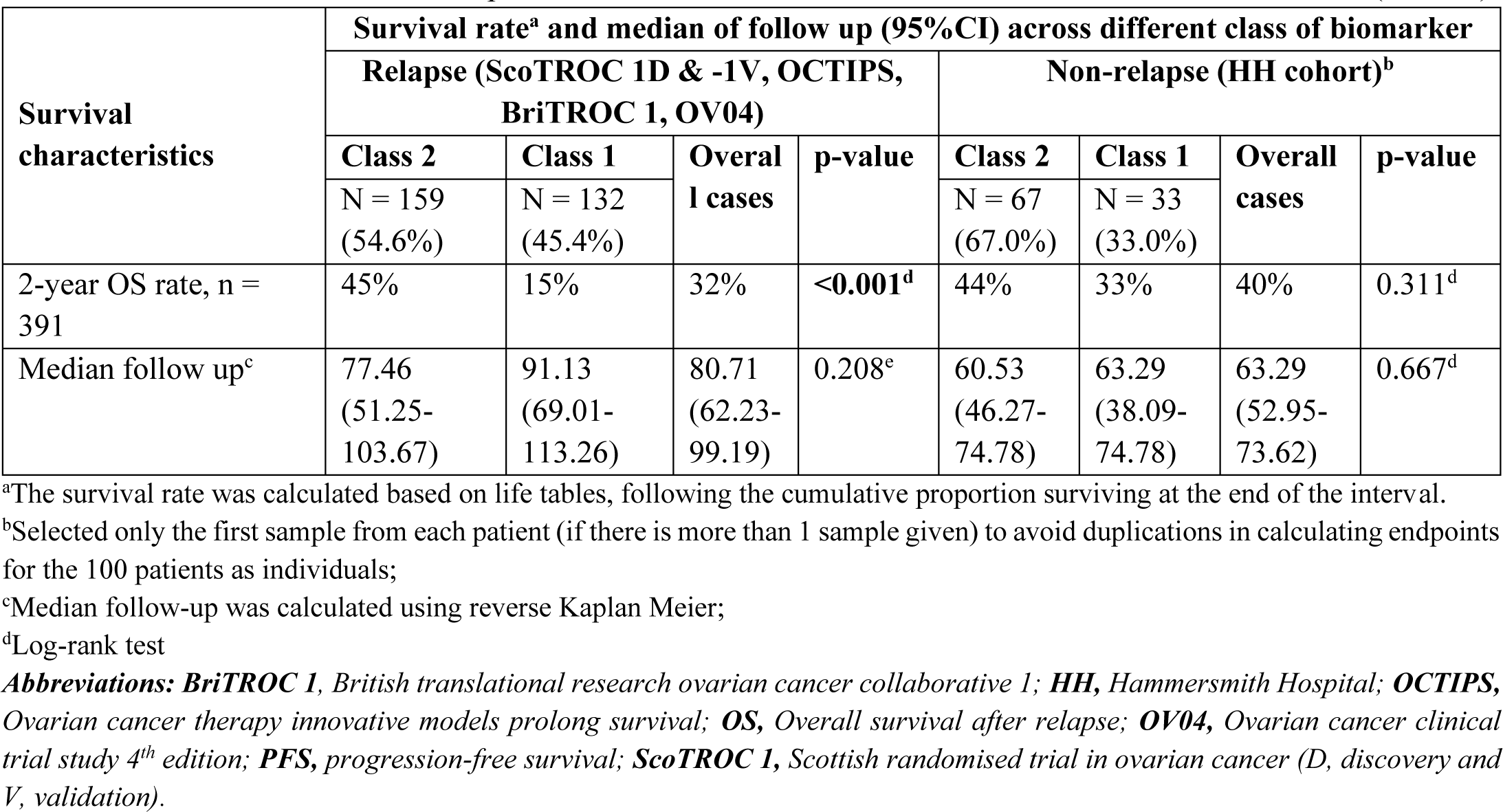
Multivariate Cox regression analysis associating overall survival after relapse with biomarker class, adjusted by clinical covariates in the Hammersmith Hospital cohort (n = 100; only complete data variables).

Looking into different cycles (cycles 3 & 4 and cycle 6) in 153 samples of the HH cohort, PLAT-M8 methylation markers showed no significant prognostic value for OS with aHR 1.34, 0.79-2.27, p=0.285 and aHR 1.11, 0.53-2.76, p=0.656, respectively (**Figure 4A, 4B**). Meta-cohort analysis (**Figure 4C**) across first-line chemotherapy patients confirmed Class 1 of PLAT-M8 did not predict a poorer prognosis. Examining methylation differences between Class 1 and Class 2 at specific CpG sites (**Figure S2**), variations were noted in HH cycles 3 & 4 and cycle 6, but these differences were not consistent with datasets from relapsed cases.

**Figure 4.**
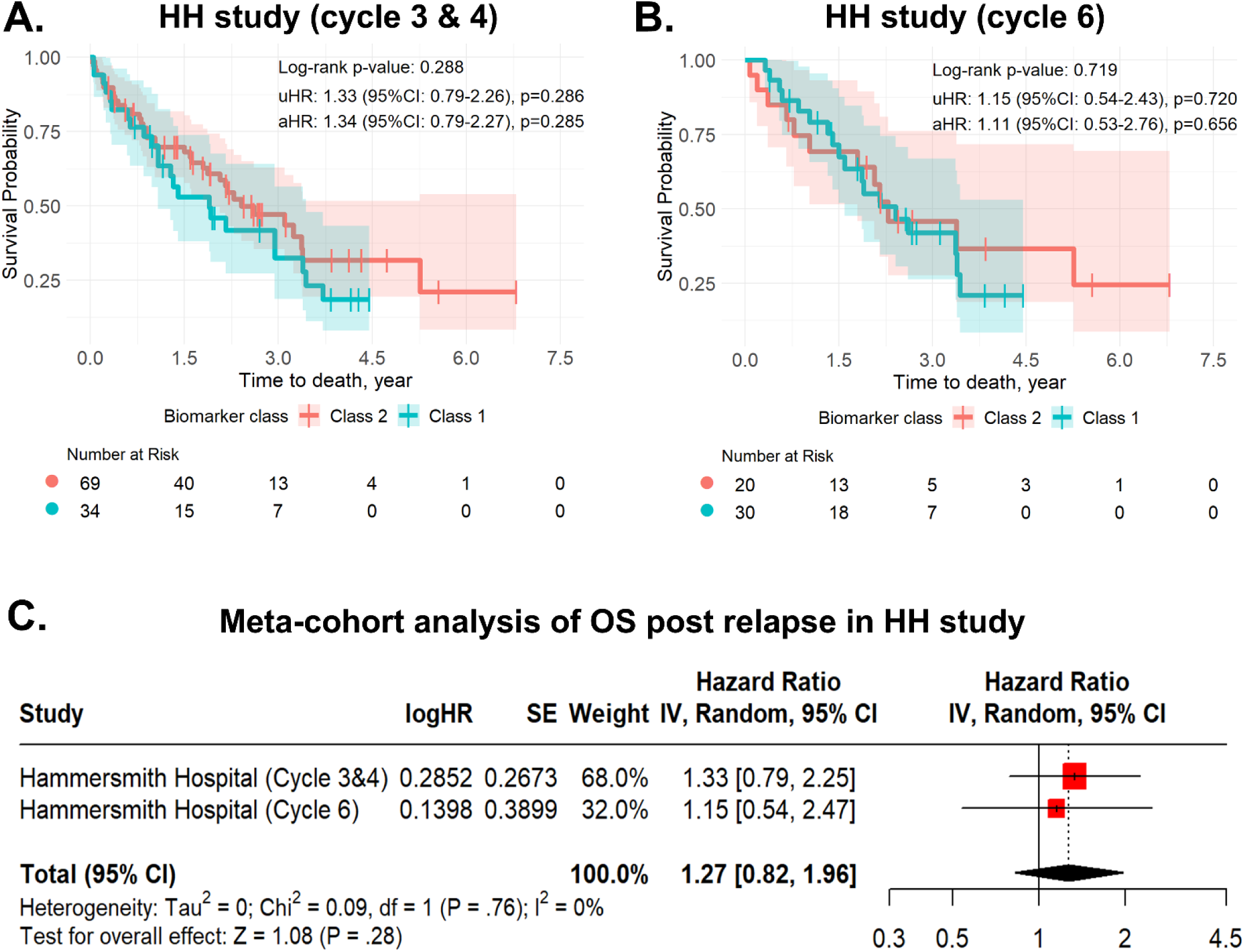
PLAT-M8 validation in Hammersmith Hospital (HH) cohort patients reveals that methylation markers are not prognostic for ovarian cancer survival during first-line chemotherapy (carboplatin + paclitaxel) before the relapse timepoint. A total of 153 blood samples from 100 patients were Analysed, focusing on cycle-specific data. In HH cycles 3&4 (n = 16, Class 1 in blue; n = 31, Class 2 in red) and HH cycle 6 (n = 25, Class 1 in blue; n = 32, Class 2 in red), Kaplan Meier (KM) curves display outcomes comparing relapse methylation Class 1 versus Class 2 (Ref). In HH cycles 3&4, both **(A)** overall survival (OS) with an adjusted hazard ratio (aHR) 1.34 (95%CI: 0.79-2.27, p = 0.285), log-rank p = 0.288 did not show significant differences between Class 1 and Class 2 of PLAT-M8. **(B)** This insignificant difference also occurs in HH cycle 6, both OS with aHR 1.11 (95%CI: 0.53-2.76, p = 0.656), log-rank p = 0.719. However, there is a tendency that Class 1 might have worse survival. All adjustments of the hazard ratio were made for the covariates of age at diagnosis, cancer stage, histology, and progression-free survival (PFS). **(C)** In the meta-cohort, considering heterogeneity, the OS analysis for detecting the PLAT-M8 biomarker among the non-relapse cases was not significant and combining studies, OS summary HR reduced to 2.06 (95%CI: 1.41-3.01, p < 0.01, I^2^ = 58%, p = 0.03).

### Prognostic performance of PLAT-M8

We analysed PLAT-M8’s ROC curves for mortality prediction using univariate and multivariate logistic regression (**Figure 5A** and **5B**) with blood and tissue biopsy and blood-only datasets **(Figure S4)**. In univariate logistic regression, PLAT-M8 alone showed moderate sensitivity (51.9%), good specificity (71.1%), and sufficient AUC (0.62). Combining it with clinical covariates improved AUC but remained within a sufficient range. To better understand PLAT-M8’s performance, we employed a dynamic control approach, including cumulative and incident case ROCs (**Figures S5** and **S6**). Cumulative case ROC assesses within the multivariate model maintain good discrimination in OS analysis, starting at AUC 0.768 and stabilizing at 0.759 in the third year for the multivariate Cox-ph model, better than the unadjusted model **(Figure 5C** and **5D)**. In this research, we conducted a sensitivity analysis on this biomarker at different cancer progression stages which indicates that PLAT-M8 is more predictive of survival after relapse in partially sensitive (PFS/PFI 6-12 months) and sensitive (PFS/PFI >12 months) patients. Additionally, our sensitivity analysis based on histological type showed that PLAT-M8 is effective for both serous and non-serous cancer. However, we could not conduct a detailed analysis of HGSOC, the most lethal OC subtype, due to a lack of histological grade information **(Table 5)**.

**Figure 5.**
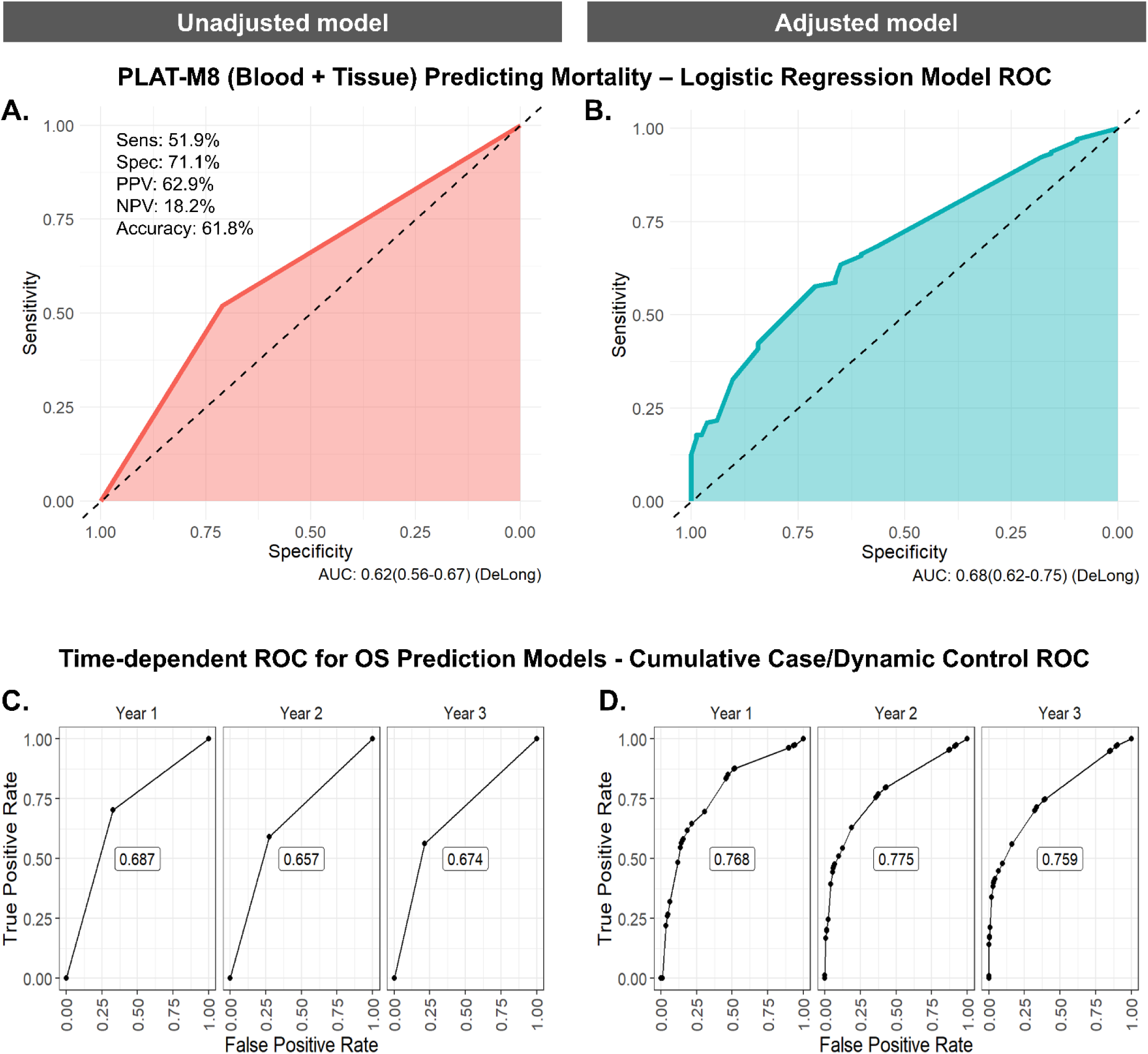
Assessing the prognostic performance of PLAT-M8 class 1 vs. class 2 (reference) in blood and tumour tissue DNA samples to predict mortality and time-dependent survival among relapsed cases (n = 291). (A) Using a univariate logistic regression model to predict mortality, PLAT-M8 alone has a sensitivity of 51.9%, specificity of 71.1%, a positive predictive value (PPV) of 62.9%, a negative predictive value (NPV) of 18.2%, and an accuracy of 61.8% with an AUC of 0.62. (B) Using a multivariate logistic regression model involving age at relapse, FIGO stage, histological type of tumour, and PFS time, PLAT-M8 may predict mortality with an improved AUC of 0.68. (C) Using a univariate Cox-regression model to assess time-dependent overall survival (OS) prediction, the performance of PLAT-M8 in predicting the cumulative incidence (events) risk over six years demonstrates its optimal discriminative value in the initial year, registering an AUC of 0.687, and subsequently decreasing to 0.674 by the third year. D) After adjustment involving age at relapse, FIGO stage, histological type of tumour, and PFS time, PLAT-M8 improves its discriminative value in the first year with an AUC of 0.768, decreasing over time to 0.759 in the third year.

**Table 5.** Sensitivity analysis of class 1 compared with class 2 of PLAT-M8 in different progression times and histological subtypes in predicting survival after first relapse.

| Subgroups | Number at risk |  | Log-rank<br>p-value | uHR (95%CI) | Coxph<br>p-value | aHR (95%CI) | Coxph<br>p-value |
| --- | --- | --- | --- | --- | --- | --- | --- |
|  | Class<br>2 | Class<br>1 |  |  |  |  |  |
| Progression time |  |  |  |  |  |  |  |
| PFS < 6 months | 5 | 20 | 0.317 | 1.87 (0.54-6.49) | 0.325 | 1.93 (0.49-7.61) | 0.350 |
| PFS 6-12 months | 30 | 56 | 0.005 | 2.08 (1.24-3.49) | **0.006** | 2.63 (1.46-4.75) | **0.001** |
| PFS >12 months | 124 | 56 | 0.008 | 1.67 (1.13-2.45) | **0.009** | 1.61 (1.08-2.40) | **0.019** |
| Histological subtype |  |  |  |  |  |  |  |
| Serous carcinoma | 119 | 87 | <0.001 | 2.12 (1.51-2.98) | <0.001 | 1.81 (1.27-2.58) | 0.001 |
| Non-serous carcinoma | 40 | 45 | <0.001 | 2.56 (1.56-4.19) | <0.001 | 2.19 (1.21-3.95) | 0.009 |

**Abbreviation:** **95%CI**, 95% confidence interval; **aHR**, adjusted hazard ratio; **PFS**, progression-free survival; **uHR**, unadjusted hazard ratio.

## Discussion

### Summary of findings

Analysing tumours at relapse is crucial to understand drug resistance and its impact on survival, but obtaining samples from relapsed patients is challenging. Consequently, plasma and serum markers, which reflect tumour DNA changes, are emerging as valuable prognostic tools.^36^ DNA methylation in plasma, requiring only small amounts, shows high specificity, making it ideal for analysis. This study validates the prognostic role of the methylation-based biomarker “PLAT-M8” in OC patients at relapse, with strong predictive value for survival risk in the first year and sustained performance over time. PLAT-M8, particularly Class 1 predicted poorer prognosis in the independent validation cohort (BriTROC 1 and OV04) but not in HH samples taken during first-line chemotherapy (pre-relapse). Significant correlations with OS were confirmed in five cohorts, and a meta-cohort analysis reinforced PLAT-M8’s reliability. This biomarker was associated with platinum sensitivity and RECIST response, but not with CA-125 response. Class 1, marked by increased hypomethylation, was associated with older patients at advanced stages, treatment resistance, higher RD, poor treatment response, and worse prognosis. These findings are consistent with an ovarian CCC study that linked DNA methylation clusters to disease features, immune pathway gene expression, and survival outcomes.^37^

### PLAT-M8 in comparison with clinicopathological characteristics and mortality status

The median age of diagnosis in our five cohorts of relapsed cases was 61 years, which is consistent with global studies.^38^ The median PFS was 16.57 months, slightly shorter than the 18-24 months reported in the literature.^39^ In our study, the age at relapse was 62 years, which was higher than that of a previous German trial (59.1 years).^38^ The age at diagnosis and relapse did not show a correlation with mortality, possibly due to different cutoffs used in the literature.^40–42^ Despite ongoing debates about the age cutoff for the elderly, the lack of a significant association between age and prognosis highlights the importance of objective assessments in treatment decisions, such as epigenetic analysis.^41^ In our study, age was found to be associated with the epigenetic signature, indicating age-related epigenetic alterations in tumorigenesis.^43^ On bivariate analysis, age at diagnosis, but not at relapse, was related to PLAT-M8. This discrepancy suggests that ageing may contribute to observed epigenetic changes long before recurrence. Our previous study suggested that age differences between presentation and relapse within a shorter time frame (<3 years) were unlikely to cause additional methylation changes.^44^ In our multivariate analysis of predictors of PLAT-M8 and mortality, we chose to use age at relapse for practical reasons during hospital visits. Epigenetic ageing may lead to cancer by accumulating random alterations that affect tumour suppressors, with accelerated epigenetic age correlating with an increased risk of cancer.^43^ Age-related changes in the tumour microenvironment, immunosenescence, stochastic DNA methylation alterations, and shifts in cellular composition collectively contribute to epigenetic variability during ageing, linking age and epigenetic dysregulation to cancer development, chemoresistance, and OS.^45^

Clinical staging significantly affects PLAT-M8, with earlier stages showing a lower relapse frequency (11%) compared to advanced stages (89%). This is consistent with evidence of recurrence in 10-50% of early-stage cases^46^ and over 80% in advanced stages.^47^ The FIGO stage is correlated with recurrence, reflecting the aggressiveness of the disease and influencing the complexity of surgery or chemotherapy, thereby increasing the likelihood of RD.^48^ The stage of the disease is associated with DNA methylation changes linked to the progression of OC.^49^ PLAT-M8 Class 1 (hypomethylation) is associated with later stages, while Class 2 (hypermethylation) is associated with earlier stages. However, the significance of the advanced stage diminishes after adjusting for other variables. Generally, abnormal DNA methylation is an early event in tumour initiation, with elevated levels of hypomethylation indicating a poor prognosis. Hypomethylation is linked to chromosome instability, increased aggressiveness, and decreased survival in OCs.^50^ Further analysis revealed that FIGO staging is an independent prognostic factor for relapsed OC, affecting ^5^ FIGO staging reflects the aggressiveness of the disease at initial presentation and relapse, potentially increasing the likelihood of the disease spreading beyond the ovary, which affects survival^51^

Despite DNA methylation’s role in epithelial OC development^52^, our analysis found no significant link between histological subtype or tumour grade and PLAT-M8. Unequal proportions in tumour histology and grade may have influenced these findings, indicating the need for a more balanced analysis. CpG methylation progressively accumulates in OC, with tumour-specific patterns of aberrant methylation.^53^ Global methylation patterns in EOC include hypermethylation of promoters and hypomethylation of repetitive DNA sequences, with histotype-specific hypermethylation reflecting differences in carcinogenic processes, immune pathways, or precursor tissue.^37,54^ Nevertheless, when comparing histological types and mortality in relapsed OC patients, we found that non-serous carcinomas—including adenocarcinoma, papillary adenocarcinoma, mucinous adenocarcinoma, endometrioid carcinoma, and CCC—demonstrated poorer survival compared to serous carcinoma. Histotype influences OC survival by affecting treatment responsiveness.^55^ Non-serous histotypes like mucinous, clear cell and carcinosarcoma respond poorly to chemotherapy and have limited targeted therapy options.^56,57^ Another study also showed that CCC is associated with a higher rate of progressive disease, and mucinous cancer shows lower response rates to first-line platinum.^58^ Despite some non-serous histotypes indicating better long-term survival, distant-stage mucinous, clear cell, and carcinosarcoma have comparable or worse 10-year survival than advanced-stage HGSOC.^55^ Early-stage diagnoses contribute to favourable outcomes for mucinous and clear cell histotypes^59^, but advanced-stage cases may have poor outcomes.^60^ In our cohorts, the poorer prognosis in non-serous cases is logical, as almost 90% of patients were at an advanced stage and included in the relapsed group.

In terms of treatment, our study found a significant correlation between second-line chemotherapy, specifically carboplatin and paclitaxel, and mortality in relapsed OC patients. Carboplatin monotherapy demonstrated a longer median PFS of approximately 3 months compared to other treatment regimens. When combined with PLAT-M8, patients in Class 2 who received carboplatin alone experienced improved PFS and OS. Data from the 1985 OCTIPS study and the 1998 ScoTROC 1 trial support these findings, leading to the 2000 recommendation to combine paclitaxel with platinum.^61,62^ Despite the overall effectiveness of chemotherapy, survival benefits in carboplatin combinations may be attributed to a lower incidence of adverse events.^63^ Patients in Class 2 consistently exhibited better prognoses, with single-agent platinum treatments potentially offering improved survival due to reduced adverse events.^61^ Multidrug combinations, particularly those involving platinum and taxanes, can result in cumulative toxicities that impact treatment adherence and patient survival.^64,65^ A meta-analysis underscores the importance of personalised approaches that consider patient factors and treatment tolerability.^64^ Adding non-cross-resistant cytotoxic drugs to carboplatin and paclitaxel aims to boost treatment efficacy but is limited by cumulative side effects, cardiotoxicity, neuropathy risks, and no clear survival benefits.^65^

In OC, surgical outcomes, specifically the status of RD after debulking surgery, are a strong prognostic factor for OS ^66,67^ and PFS.^67,68^ RD indicates adverse tumour biology, correlating with severe dissemination and progression^66^, affecting both primary and relapse cases.^69,70^ In our analysis, although the type of debulking surgery does not directly correlate with methylation status, RD resulting from surgery is associated with mortality. Attaining optimal debulking is crucial for maximizing survival benefits, increasing median OS by 5.5% and maximises cytoreduction rates by 10%.^66^ Unfortunately, in our study, 60.1% of patients had RD, which possibly contributed to recurrence after initial treatment.^71^ Additionally, bivariate analysis showed that RD resulting from surgery was associated with Class 1 of PLAT-M8, indicating its relation to methylation changes. Previous studies have linked suboptimal surgical outcomes to epigenetic changes, specifically hypomethylation of *SPARC*. This hypomethylation leads to increased gene expression and invasiveness.^72^ Moreover, RD may represent cells with unique methylation patterns, contributing to treatment resistance.^73^ Accordingly, it is proposed that PLAT-M8 has potential as a biomarker for detecting and monitoring RD. It can provide insights into treatment response and changes in DNA methylation patterns induced by therapies like chemotherapy and radiotherapy.

Further analysis of parameters related to recurrence revealed that PLAT-M8 and RECIST response intersects with epigenetic methylation in OC. While previous studies have explored the relationship between ctDNA progression and RECIST response, they have not specifically investigated the role of different methylation patterns in this process.^74^ Therefore, the novel PLAT-M8, which includes aberrant methylation information, offers new insights into this correlation. Elucidating the link between RECIST and PLAT-M8 can enhance the personalised management of ovarian cancer by integrating anatomical changes with epigenetic influences. Previous studies have used RECIST to measure treatment response with methylation biomarkers in cancers such as breast^75^ and cervical cancer.^76^ In metastatic colorectal cancer, RECIST response correlated with hypermethylated *TFAP2E*, linked to chemoresistance.^77^ Similarly, hypermethylated neuropeptide Y was an early biomarker outperforming RECIST in predicting PFS.^78^ Our study suggests that PLAT-M8, associated with RECIST and potentially better at predicting recurrence, might surpass conventional imaging markers due to its epigenetic basis, which reflects tumour behaviour and predicts OS more effectively.^79^

Currently, there is no standard prognostic indicator for OC, especially in relapsed cases. CA-125, a widely used protein biomarker, has controversial sensitivity and specificity.^80^ DNA methylation markers offer a stable and specific alternative, enabling personalised treatment plans based on the overall DNA methylation profile of ovarian tumours. CA-125’s clinical use is limited by its inadequate sensitivity (50-62%) and uncertain prognostic role in relapsed OC.^81^ It does not significantly correlate with mortality status and lacks conclusive evidence for predicting OS.^82^ In our research, PLAT-M8 at relapse does not statistically correlate with CA-125. This aligns with CA-125’s limited prognostic role in relapsed ovarian cancer and its minimal significance in recurrence-related factors. ^83^ The prognostic value of CA-125 varies in platinum-sensitive and --resistant OC, and it shows limited predictability for complete tumour resection.^84^ We also analysed another clinical factor called PFI, which is considered during recurrence. Our aim was to replace it because it inaccurately classifies recurrences based on time.^85,86^ Although we found that PFI was linked to mortality status, PFS, and OS, it was not consistent as a prognostic factor for relapsed OS patients in previous research.^87^ On the other hand, PLAT-M8 is believed to be a more accurate prognostic biomarker that reflects the sensitivity level to platinum. PFI, lacking molecular insights, does not adequately represent platinum resistance and fails to differentiate platinum sensitivity in different clinicopathological contexts.^88,89^

Our analysis shows that PLAT-M8 is superior, correlating with PFS in both univariate and multivariate analyses across relapsed cases. Research has linked PFS with methylation, identifying 112 loci associated with shorter PFS after chemotherapy^90^ and an 11-gene panel linked to the *Bmi-1* pathway connecting shortened PFS with distant metastasis.^91^ In our analysis, we observed that shorter PFS is a strong predictor of mortality after relapse. This is an important finding as previous studies have also used time to recurrence/PFS as a factor to predict survival after recurrence in advanced-stage HGSOC.^92^ Time to recurrence or PFS, a phenotype that reflects cancer genome and epigenome characteristics.^92^

### PLAT-M8 as an epigenetic-based prognostic biomarker

PLAT-M8 demonstrates strong prognostic value for survival after relapse, even without information regarding secondary cytoreductive surgery. PLAT-M8 Class 1, characterised by hypomethylation, aligns with studies that link hypomethylation in tumour-initiating cells to poor OC prognosis, specifically *ATG4A* and *HIST1H2BN* hypomethylation.^93^ During validation, PLAT-M8 significantly predicted OS post-relapse in the BriTROC 1 and OV04 datasets, as well as five additional datasets, confirming its effectiveness across multiple sources. PLAT-M8 is effective only during relapse, not before or during initial chemotherapy, indicating it reflects platinum sensitivity post-relapse rather than pre-existing chemoresistance.

For sensitivity analysis, we derived platinum sensitivity categories (i.e., PFI) from cancer progression stages (i.e., PFS), despite their different starting points: PFI starts from the last platinum treatment to progression/recurrence, while PFS begins at initial surgery.^94^ Mankoo et al.^95^ found a direct correlation between PFS and PFI outcomes, justifying the use of PFS to gauge responsiveness to platinum when PFI data is unavailable. One key finding is that PLAT-M8 was a good predictor of survival after relapse in partially sensitive cases, which is crucial for deciding on further platinum therapy. Our analysis also confirmed PLAT-M8’s effectiveness for both serous and non-serous cancers. However, a detailed analysis of HGSOC was limited by a lack of histological grade data. Nonetheless, since 70-90% of serous carcinoma is classified as HGSOC, which frequently relapses, PLAT-M8 can be considered a robust prognostic biomarker across major OC subtypes.^96^

Additionally, PLAT-M8 was validated as a prognostic factor for relapsed OC across various second-line chemotherapy regimens using the BriTROC 1 and OV04 datasets. Despite factors like PFI, patient status, and toxicity guiding treatment, there is a lack of DNA-methylation-based biomarkers in decision-making.^97^ In our study, platinum monotherapy was effective for Class 2 biomarkers (mostly platinum-sensitive), with survival rates comparable to Class 2 patients treated with other therapies with/without platinum. For Class 1 patients (mostly platinum-resistant), rechallenging with other therapies (paclitaxel/docetaxel, liposomal doxorubicin, and topotecan) with/without platinum combinations showed similar or better outcomes than platinum alone, consistent with previous findings.^98,99^ Platinum-based combinations, especially in late recurrent disease, are known to provide better outcomes, suggesting that Class 1 patients may still benefit from them.^100^ This indicates that resistance based on PFI might exclude patients from benefiting from other therapies with/without platinum-based combinations. On the other hand, PLAT-M8 can still tailor treatments to help Class 1 patients benefit from these therapies. Nevertheless, platinum-only treatment for Class 1 patients should not be advised as it worsens survival outcomes. Interestingly, Class 2 patients receiving multiple second-line combinations had poorer survival than Class 1, possibly due to unpredictable toxicity or treatment discontinuation.^101^ Compliance issues with platinum combinations may extend response duration, but prolonging therapy and adding new drugs risk increased toxicity, costs, and treatment dropout without significant benefits.^101^ Larger randomised trials are needed to evaluate these strategies, with PLAT-M8 potentially serving as a predictive biomarker for optimising platinum regimens in second-line therapy for relapsed OC.

### Proposed mechanisms

PLAT-M8 involves mismatch repair, specifically *MLH1*, which indicates hypermethylation in Class 2. Mismatch repair recognises platinum-induced DNA damage, leading to hypermethylation. In Class 2, hypermethylation after platinum is caused by platinum adducts that result in replication stalling in MLH1-expressing cells, leading to cell death.^14^ This hypermethylation contributes to improved patient survival and establishes a link between DNA methylation, PLAT-M8, platinum-induced damage, and its related oncogenes silencing. Dysfunctional repair systems enable cells to bypass platinum-related issues, promoting resistance.^102^ PLAT-M8, consisting of 8 CpGs, is associated with genes (*ZNF385D, ZPLD1, MAD1L1, SAMD12, ARID5B, DUSP6, PPP2R5E,* and *SBNO2*) that are linked to carcinogenesis, chemoresistance, and prognosis. Although these genes have been less explored in OC, they have shown prognostic significance in other cancers. For example, *ZNF385D* and *ZPLD1* have prognostic value in liver^103^ and breast cancer^104^, respectively. *MAD1L1’*s promoter hypermethylation affects advanced OC prognosis^105^ and correlates with chemotherapy response.^106^ *SAMD12* is relevant in gastric cancer prognosis ^107^, reduced *ARID5B* expression indicates poor prognosis in OC^108^, and *DUSP6* overexpression is linked to chemotherapy resistance.^109^ *SBNO2* levels are associated with adverse prognoses in cervical cancer.^110,111^ Further experimental validation is needed to confirm their involvement by assessing the impact of methylation on gene expression.

### Strengths and limitations

This study meticulously analyses clinicopathological characteristics related to methylation-based biomarker diseases using five independent relapsed OC cohorts, including OCTIPS tissue samples. The multi-institutional design with large UK ovarian cohorts improves result generalisability. The retrospective design provides access to clinical endpoints and characteristics, and blood-based samples offer practicality and less invasiveness, benefiting personalised medicine for relapsed patients. The retrospective design limits data on treatment response, especially second-line treatment. Future research should focus on predicting second-line treatment response and more comprehensively capturing chemotherapy response and survival mechanisms. Reliance on archival DNA biospecimens complicates gene-related 8 CpGs expression analysis due to methylation changes. Additionally, using DNA from white blood cells may not fully reflect tumour DNA compared to more accurate but costly ctDNA methylation analysis.

## Conclusion

PLAT-M8, rooted in epigenetics, emerges as a potential game-changer, offering a reliable biomarker for relapsed OC and potentially preventing ineffective chemotherapy reintroduction. This validation study reveals clinicopathological features associated with different PLAT-M8 Classes. Class 1 indicates a hypomethylated signature, indicating poorer outcomes. This class presents as an older population at an advanced stage, resistant to treatment, and more progressive. Our findings support PLAT-M8’s potential as a valuable prognostic marker in OC at relapse, both in blood samples and tissue biopsies. This insight contributes to stratifying second-line treatment strategies and predicting survival. Future research will focus on PLAT-M8’s role in predicting second-line treatment response and understanding its mechanism in chemoresistance, emphasizing CpG-associated gene expression influenced by methylation. Further validation and cost-effectiveness assessments are needed to establish PLAT-M8’s clinical position relative to existing biomarkers like CA-125, RECIST, or PFI, with future comparisons among patients with different responses to second-line treatment based on RECIST criteria.

## Data Availability

All the reporting studies adhere to the REporting recommendations for tumour MARKer prognostic studies (REMARK)[33] and The Strengthening the Reporting of Observational Studies in Epidemiology (STROBE) guidelines.[34]. All data produced in the present work are contained in the manuscript.

## Authors’ Contributions

JMF served as the principal investigator for this study, secured funding, and made the decision to publish. As the guarantor, JMF assumed full responsibility for the work. JMF and MH conceptualised the research, contributed to the analysis, conducted the investigation, designed the methodology, managed project administration, utilised software, and created visualisations of the study findings. They had complete access to the literature data, conducted reviews and revisions, and drafted the paper. Additionally, JMF, MH, NM, NP, AMP, RB, JDB, and IAM curated data, provided resources, and reviewed and edited the manuscript. JMF, RB, JDB, and IAM validated all evidence analyses, while JM and IAM thoroughly supervised the study process. All mentioned authors critically revised the manuscript for important intellectual content and approved the final version for publication.

## Acknowledgements

The first author would like to acknowledge a scholarship from *Beasiswa LPDP* (the Indonesian Endowment Fund for Education) from the Ministry of Finance of the Republic of Indonesia, which provided immense support to pursue a PhD at Imperial College London under contract number 20230722299954.

## Patient consent for publication

The patient has provided consent and permission to participate in this study and for publication, with identity details concealed in accordance with the principles of the Declaration of Helsinki.

## Data sharing statement (Availability of data and material)

All relevant data and materials supporting the findings of this study are available upon request from the corresponding author.

## Figure guidelines

We confirm that our figures have complied with the journal image preparation guidelines. Also, there is no image manipulation in this study.

## SUPPLEMENTARY FIGURES

**Figure S1.**
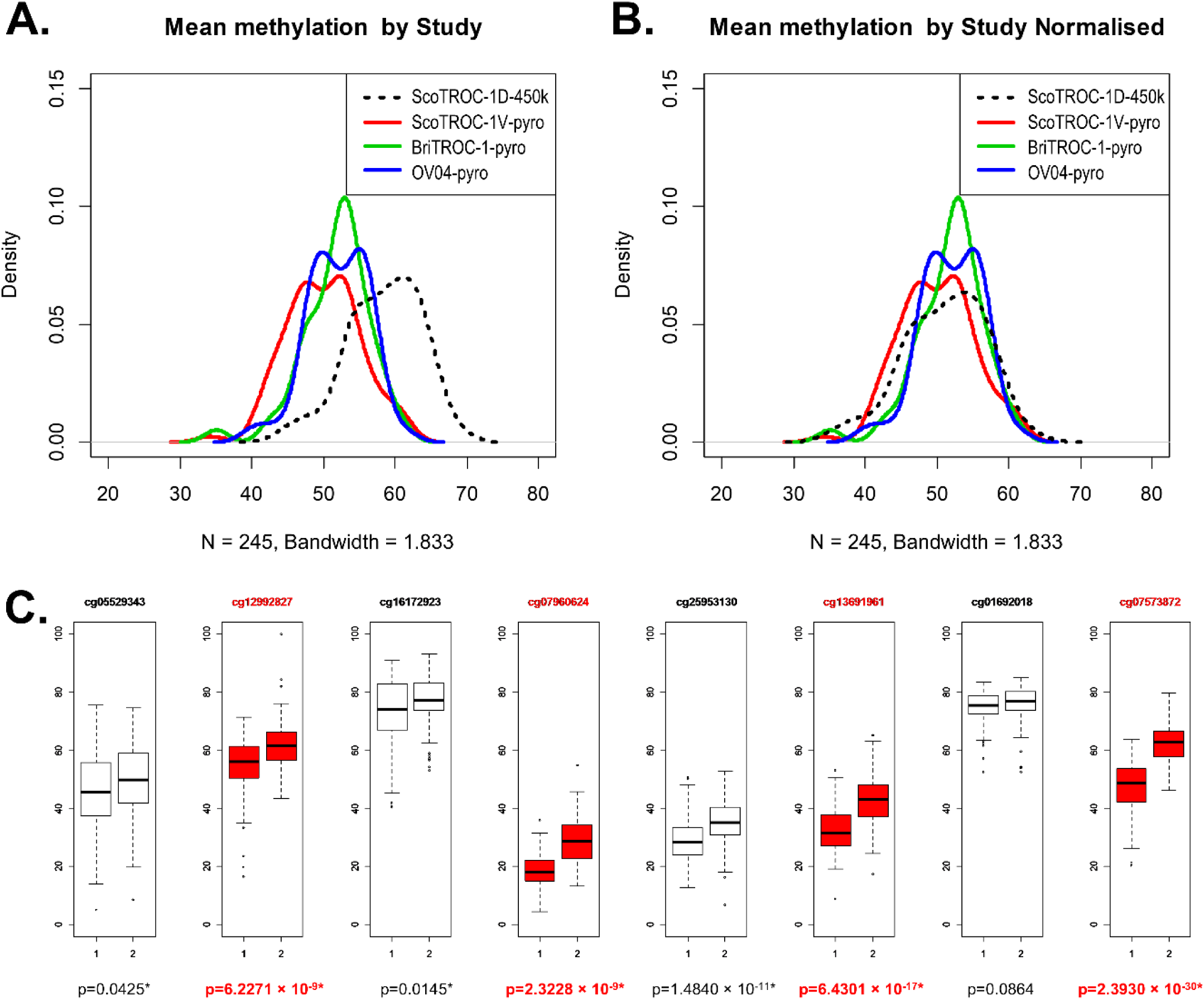
Quality control and methylation analysis of blood samples from relapsed ovarian cancer patients across four cohorts. (Note: OCTIPS study excluded due to methylation data originating from tissue biopsy, not blood.) Bandwidth: 1.833. DNA sample numbers are indicated for each cohort (ScoTROC 1-450K = 54, ScoTROC 1-pyro = 87, BriTROC 1-pyro = 47, OV04 = 57). **(A)** ScoTROC 1 shows distinct mean methylation (%) distribution due to the different methylation analysis techniques (450K methylation array was used in their discovery process and pyrosequencing was used in the validation). **(B)** The distribution of mean DNA methylation percentages following normalisation is depicted in the figure, with a black dashed line indicating the adjusted average through reference normalisation. Prior to normalisation, ScoTROC 1D-450K exhibited a median of 58.88% (IQR: 54.53-62.44%) and a mean of 58.46% (Min. 44.64%, Max. 68.96%). After normalisation, ScoTROC 1D-450K showed a median of 51.47% (IQR: 46.64-55.58%) and a mean of 51.05% (Min. 36.00%, Max. 63.01%). Other cohorts post-normalisation included ScoTROC 1V-pyro (Median 49.85%, IaQR: 46.48-50.01%, Mean 53.15%, Min. 34.18%, Max. 61.20%), BriTROC 1-pyro (Median 52.56%, IQR: 48.93-54.19%, Mean 51.81%, Min. 35.07%, Max. 60.99%), and OV04-pyro (Median 51.98%, IQR: 48.61-55.10%, Mean 52.02%, Min. 39.81%, Max. 61.55%). Most probes showed methylation levels between 30% and 70% at a density level of 0.05 to 0.1, with a prominent peak mean around 52% DNA methylation. BriTROC 1 and ScoTROC 1-450K had a single peak, whereas ScoTROC 1-pyro and OV04-pyro displayed dual peaks. The normalisation process involved centring and scaling. **(C)** All CpG sites were examined across 245 DNA samples from four cohorts. Highlighted in red are the four CpG sites selected using Elastic Net: cg12992827, cg21625271, cg07960624, and cg13691961. Hypermethylation was observed in 7 out of 8 CpG sites (Wilcoxon Test, p < 0.05, indicated by an asterisk). Each boxplot shows % methylation (Y-axis) for the n samples from the Infinium HumanMethylation450k BeadChip and pyrosequencer after normalisation for class 1 and class 2 samples (x-axis).

**Figure S2.**
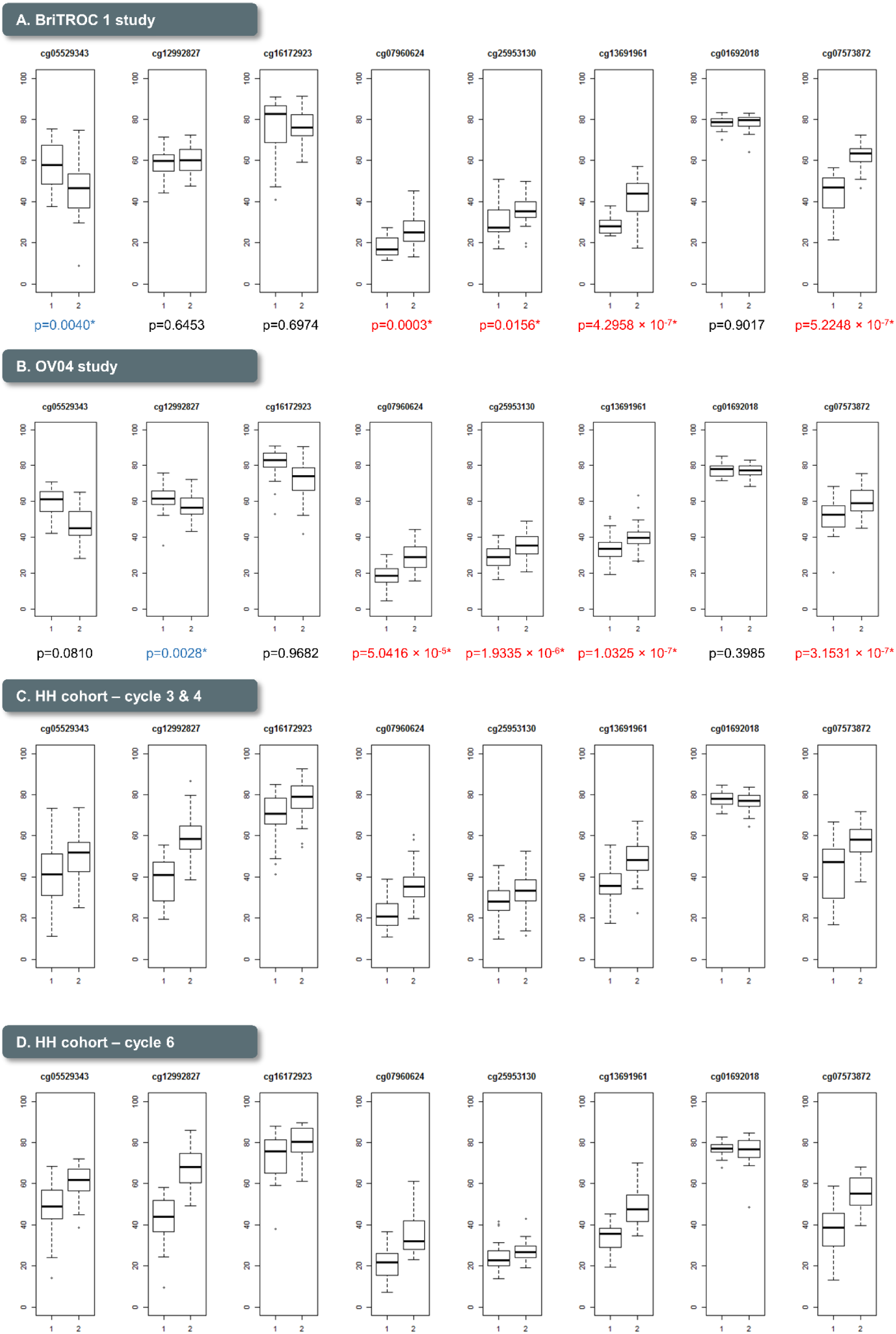
Methylation levels (%) in blood DNA from relapsed and non-relapsed ovarian cancer patients, comparing Class 1 and Class 2 of the PLAT-M8 biomarker using a pyrosequencer**. (A)** BriTROC 1 study (n = 16, Class 1; n = 31, Class 2) shows significant hypermethylation in Class 2 at 4/8 CpG sites (p < 0.05, indicated by an asterisk and red), and hypomethylation at 1/8 CpG sites (p < 0.05, indicated by an asterisk and blue). **(B)** OV04 study (n = 25, Class 1; n = 32, Class 2) reveals similar patterns with 4/8 CpG sites hypermethylated (p < 0.05, indicated by an asterisk and red) and 1/8 CpG sites hypomethylated (p < 0.05, indicated by an asterisk and blue). **(C)** Hammersmith Hospital cycle 3&4 study (n = 52, Class 1; n = 51, Class 2) does not show significant differences in methylation **(D)** Hammersmith Hospital cycle 6 study (n = 24, Class 1; n = 26, Class 2) does not show significant differences in methylation.

**Figure S3.**
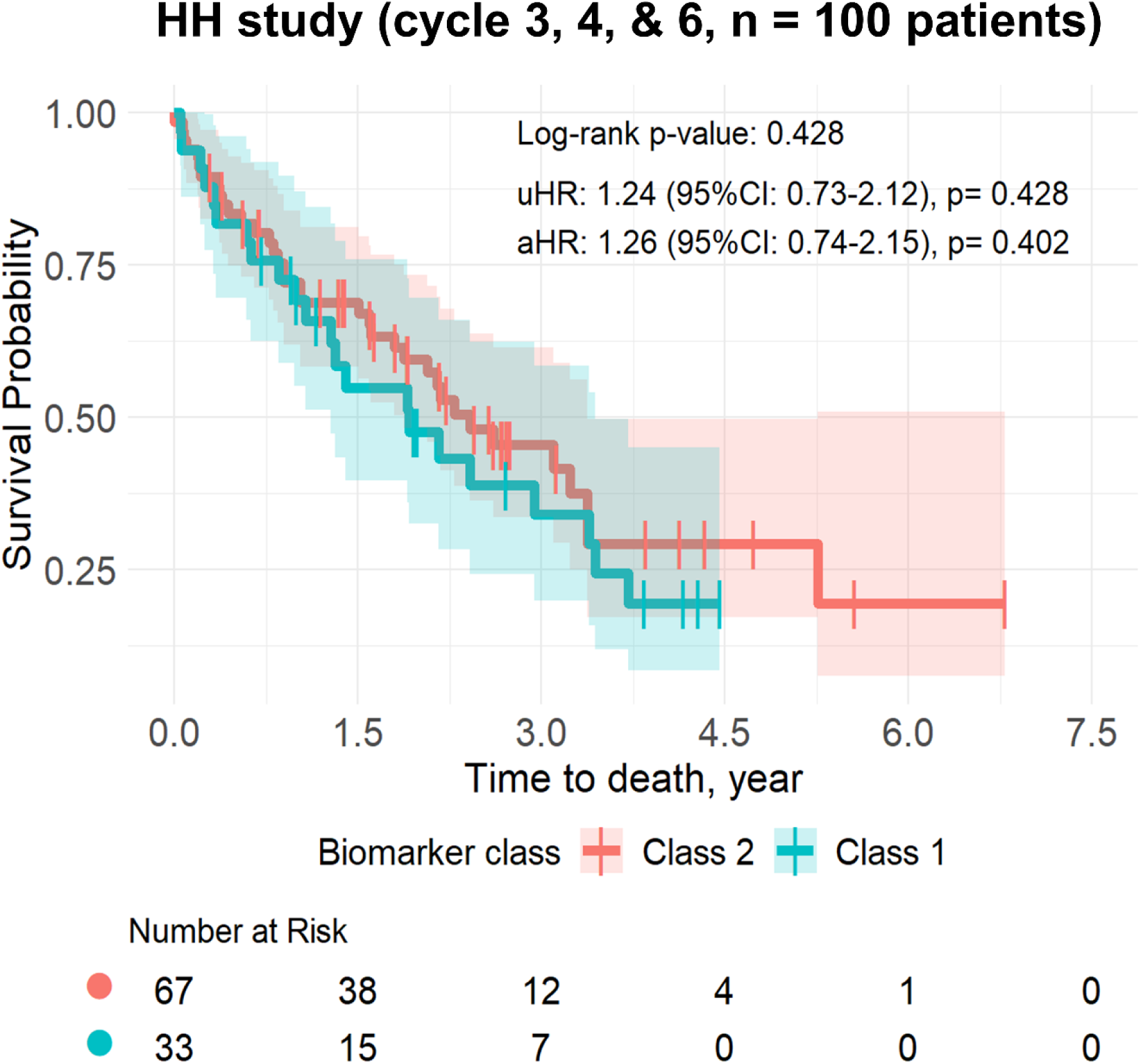
Methylation markers are not prognostic for ovarian cancer survival during first-line chemotherapy (analysis focuses on 100 patients, excluding cycle-specific data). Blood samples were collected from the Hammersmith Hospital cohort during the first-line chemotherapy (Carboplatin + Paclitaxel) treatment course. For this analysis, we selected only the initial sample from each patient to avoid duplications in calculating endpoints for the 100 patients. This analysis differs from the prior figure, which included 153 samples. Analysing 100 patients, overall survival (OS) after relapse did not show significant differences between Class 1 (n = 33) and Class 2 (n = 67) of PLAT-M8. However, there is a tendency that Class 1 might have worse survival. All adjustments were made for the covariates of age at diagnosis, cancer stage, histology, and progression-free survival (PFS).

**Figure S4.**
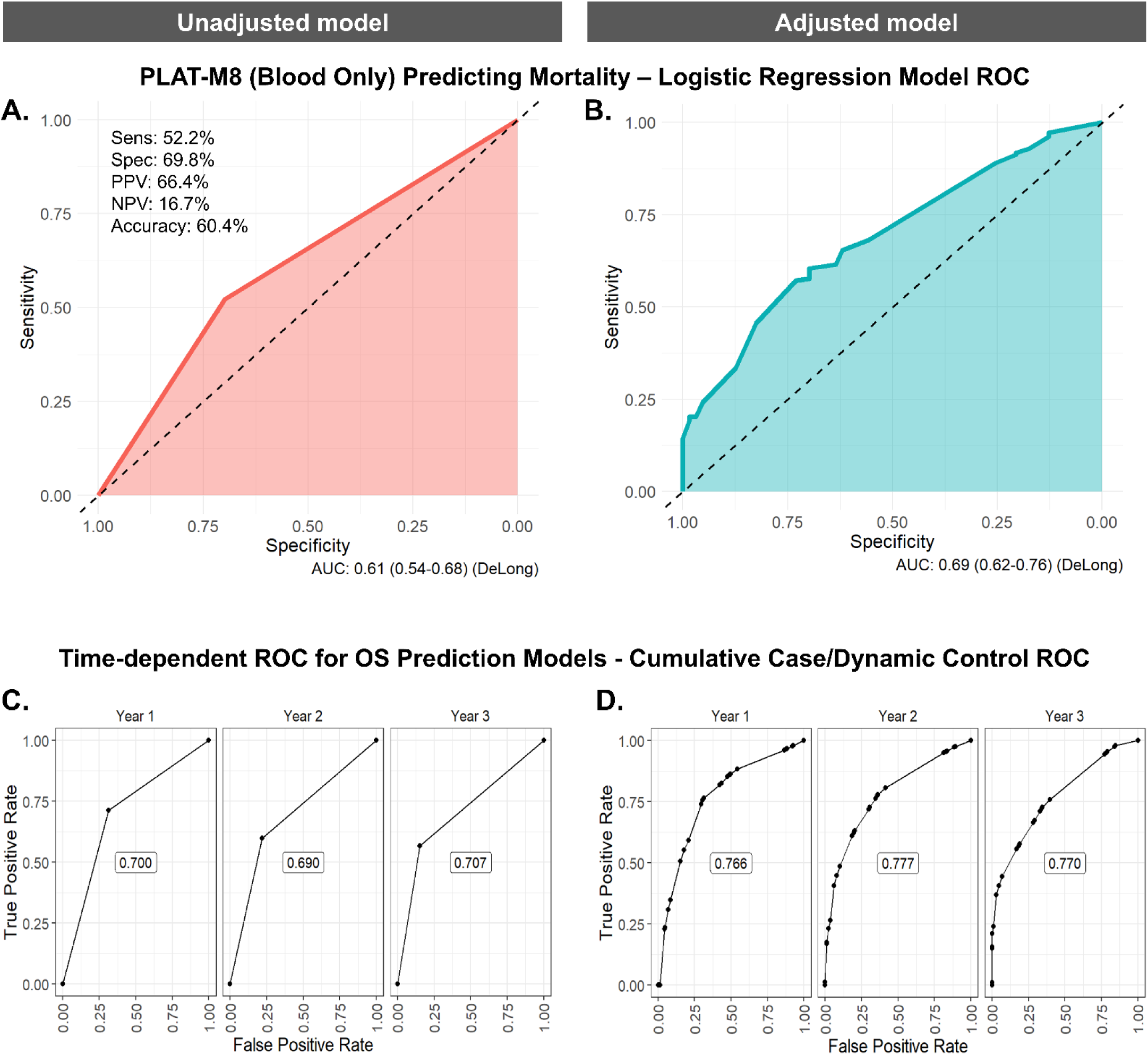
Assessing the prognostic performance of PLAT-M8 class 1 vs. class 2 (reference) in blood DNA samples to predict mortality and time-dependent survival among relapsed cases (n = 245). (**A)** Using a univariate logistic regression model to predict mortality, PLAT-M8 alone has a sensitivity of 52.2%, specificity of 69.8%, a positive predictive value (PPV) of 66.4%, a negative predictive value (NPV) of 16.7%, and an accuracy of 60.4% with an AUC of 0.61. (**B)** Using a multivariate logistic regression model involving age at relapse, FIGO stage, histological type of tumour, and PFS time, PLAT-M8 may predict mortality with an improved AUC of 0.69. (**C)** Using a univariate Cox-regression model to assess time-dependent overall survival (OS) prediction, the performance of PLAT-M8 in predicting the cumulative incidence (events) risk illustrates its superior discriminative capability over 3 years. It excels particularly in the first year, boasting an AUC of 0.700, and experiences a slight increase to 0.707 by the third year. **(D)** After adjustment, PLAT-M8 maintains a consistent discriminative value, with an AUC of 0.766 persisting from the first year to the third year.

**Figure S5.**
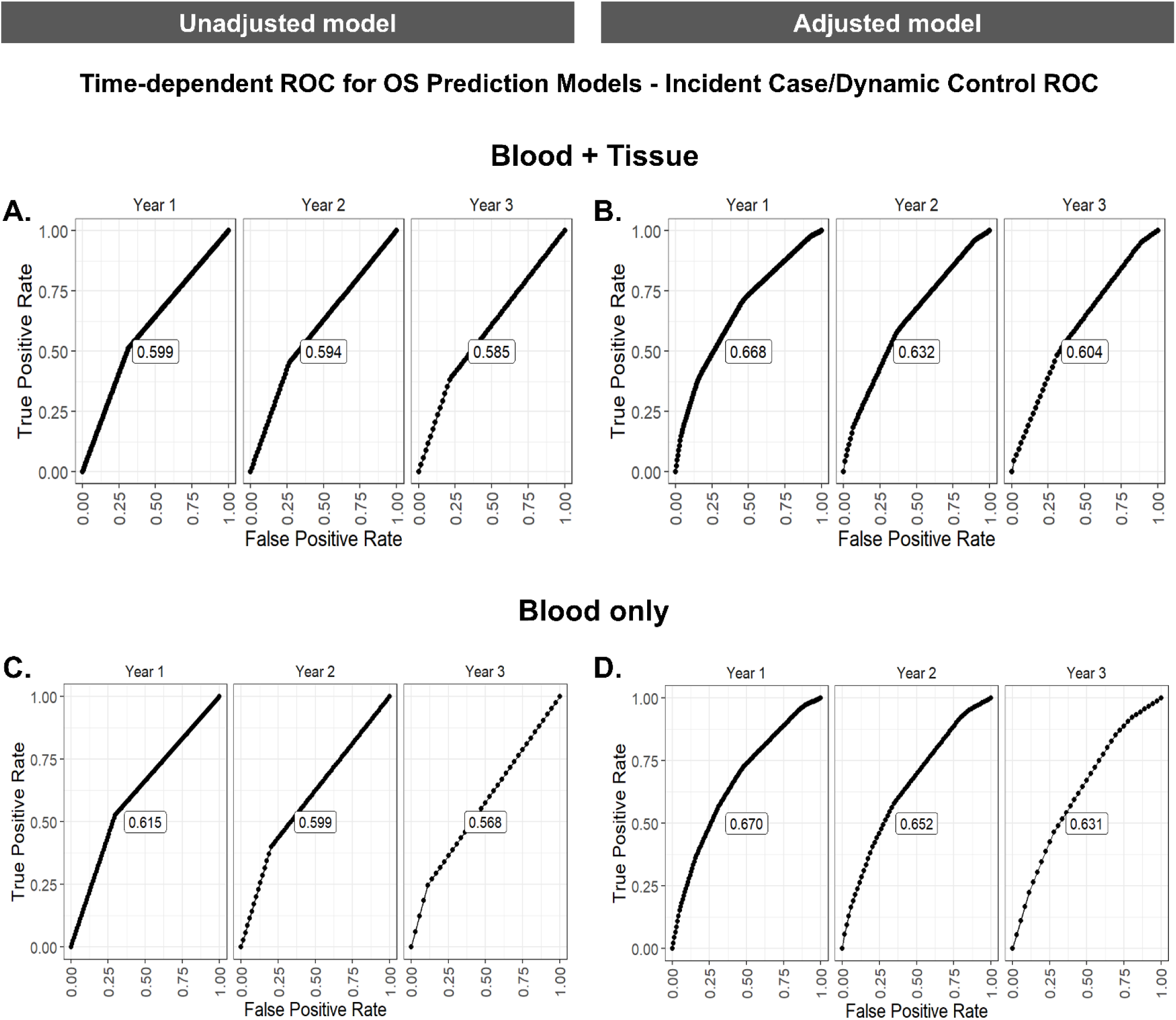
Time-dependent ROC curves for survival prediction models using blood and tissue biopsy DNA samples (n = 291) and blood DNA samples only (n = 245). **(A)** The univariate Cox-regression model for time-dependent overall survival (OS) prediction using PLAT-M8 demonstrates sufficient discriminative value, with an initial AUC of 0.599 in the first year and a slight decrease to 0.585 by the third year for predicting hazards at the 3-year mark. **(B)** After adjustment involving age at relapse, FIGO stage, histological type of tumour, and PFS time, PLAT-M8 improves its discriminative value in the first year with an AUC of 0.668, decreasing over time to 0.604 in the third year. **(C)** The univariate Cox-regression model for time-dependent OS prediction using PLAT-M8 demonstrates sufficient discriminative value, with an initial AUC of 0.615 in the first year and a slight decrease to 0.568 by the third year for predicting hazards at the 3-year mark. **(D)** After adjustment involving age at relapse, FIGO stage, histological type of tumour, and PFS time, PLAT-M8 improves its discriminative value in the first year with an AUC of 0.670, decreasing over time to 0.631 in the third year.

## SUPPLEMENTARY TABLES

**Table S1.** Distribution of PLAT-M8 classification in different datasets of cohorts, according to clustering analysis consensus (n = 391 patients, 444 samples).

| Cohorts | Biomarker status<sup>a</sup> |  |  |  | Total |  | p-value |
| --- | --- | --- | --- | --- | --- | --- | --- |
|  | Class 2 |  | Class 1 |  |  |  |  |
|  | n | % | n | % | n | % |  |
| ScoTROC 1D | 25 | 46.3 | 29 | 53.7 | 54 | 12.2 | 0.013<sup>a</sup> |
| ScoTROC 1V | 43 | 49.4 | 44 | 50.6 | 87 | 19.6 |  |
| OCTIPS | 28 | 60.9 | 18 | 39.1 | 46 | 10.4 |  |
| BriTROC 1 | 31 | 66.0 | 16 | 34.0 | 47 | 10.6 |  |
| OV04 | 32 | 56.1 | 25 | 43.9 | 57 | 12.8 |  |
| HH sample on cycles 3&4<sup>b</sup> | 69 | 67.0 | 34 | 33.0 | 103 | 23.2 |  |
| HH sample on cycle 6<sup>b</sup> | 20 | 40.0 | 30 | 60.0 | 50 | 11.3 |  |
| Total relapse cases | 159 | 54.6 | 132 | 45.4 | 291 |  |  |
| Total non-relapse samples | 89 | 58.2 | 64 | 41.8 | 153 |  |  |
| Total overall | 248 | 55.9 | 196 | 44.1 | 444 |  |  |

The percentage ‘%’ values in the biomarker status column represent row percentages, meanwhile in total column represent column percentages. <sup>a</sup>Chi-square test. <sup>b</sup>When the analysis focuses on 100 patients, excluding cycle-specific data, the number of patients in each biomarker status is Class 1 (n = 33) and Class 2 (n = 67)

**Table S2.**
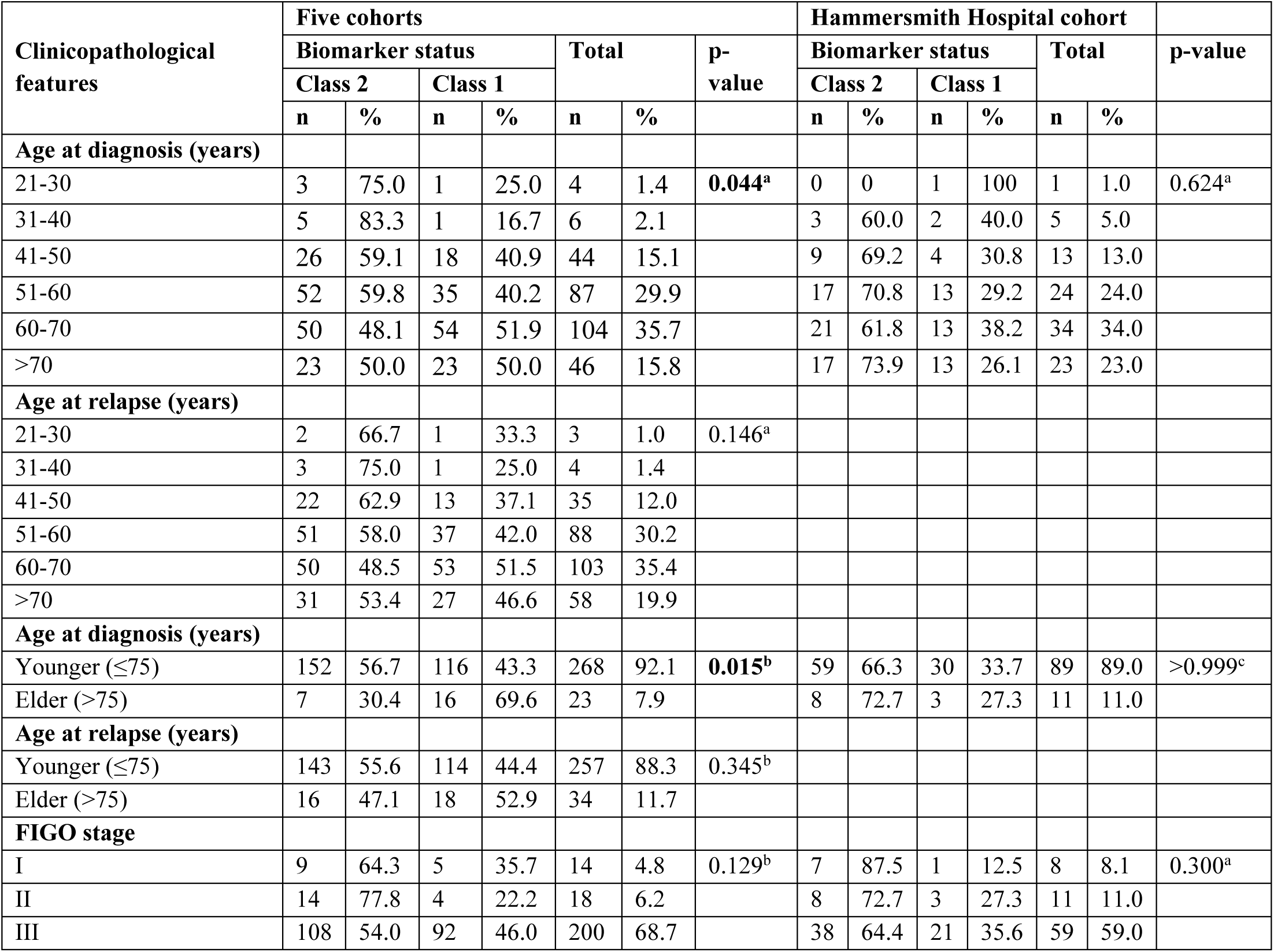
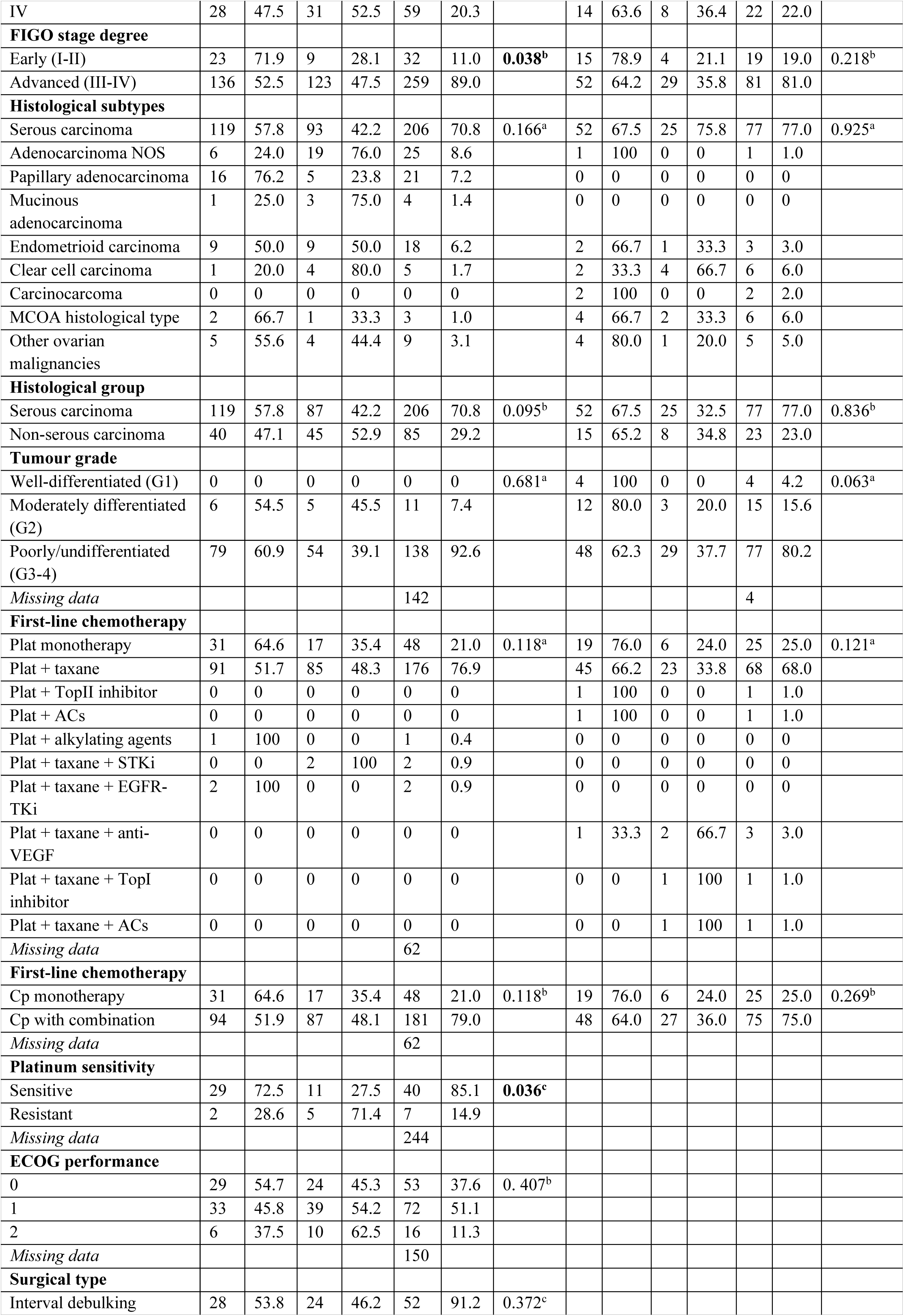
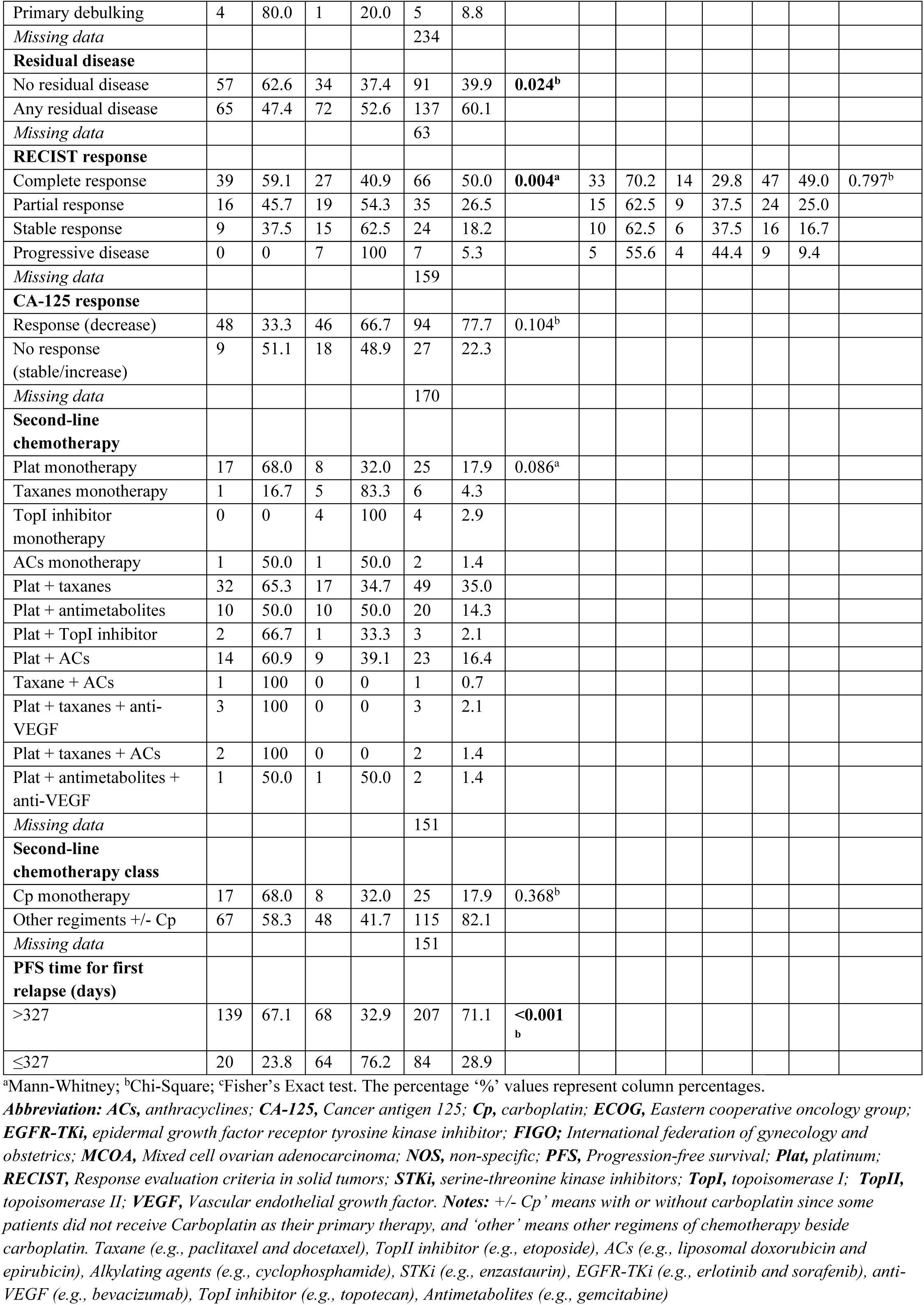
Clinicopathological features in five cohorts with relapsed ovarian cancer cases (n = 291) and at Hammersmith Hospital with non-relapsed cases (n = 100) in relation to biomarker status (negative and positive).

**Table S3.**
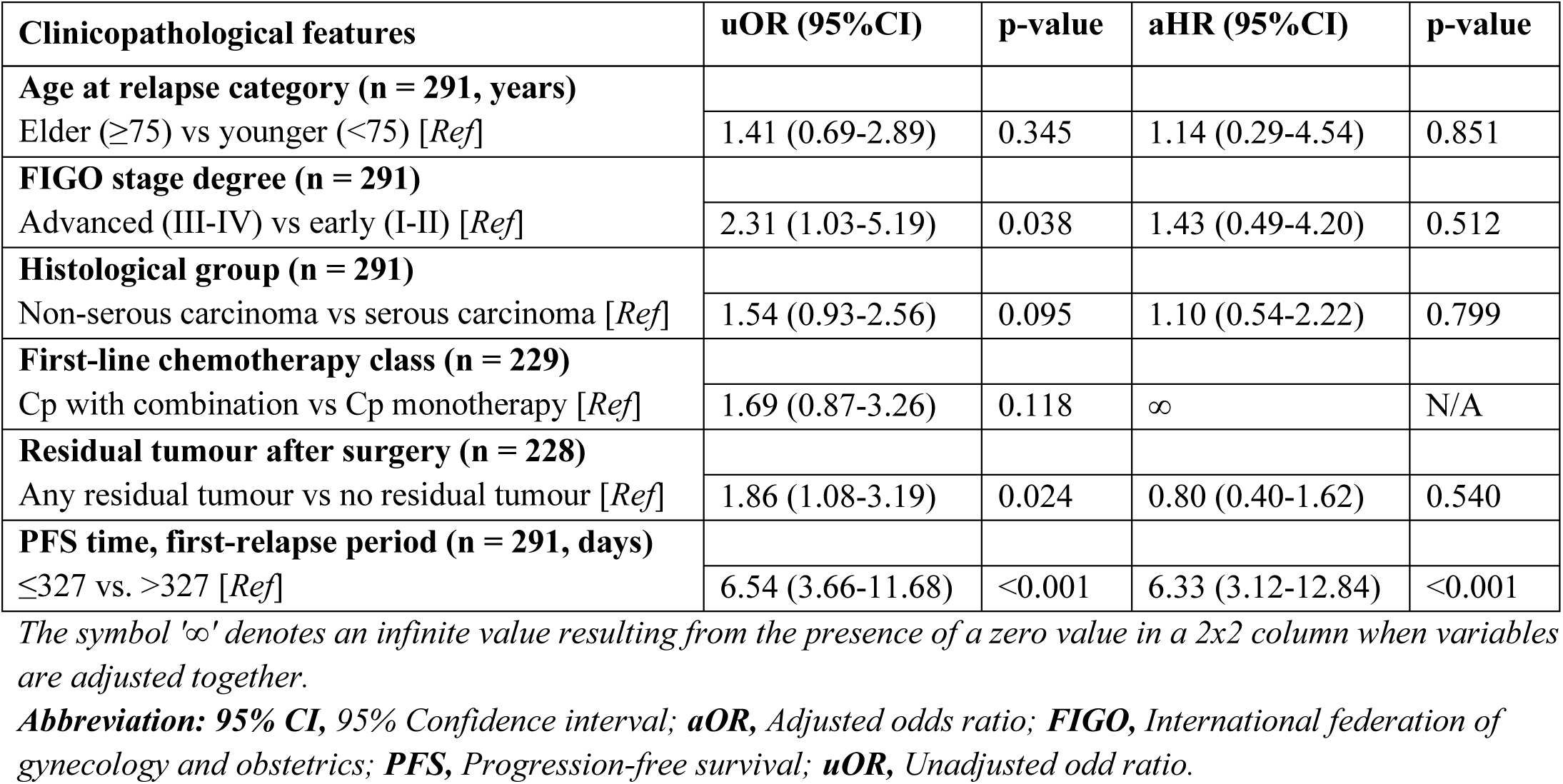
Multivariate logistic regression analysis of factors associated with Class-1 biomarker (negative epigenetic changes and poor outcome) using available data in cohorts (combined total n = 180 patients).

**Table S4.**
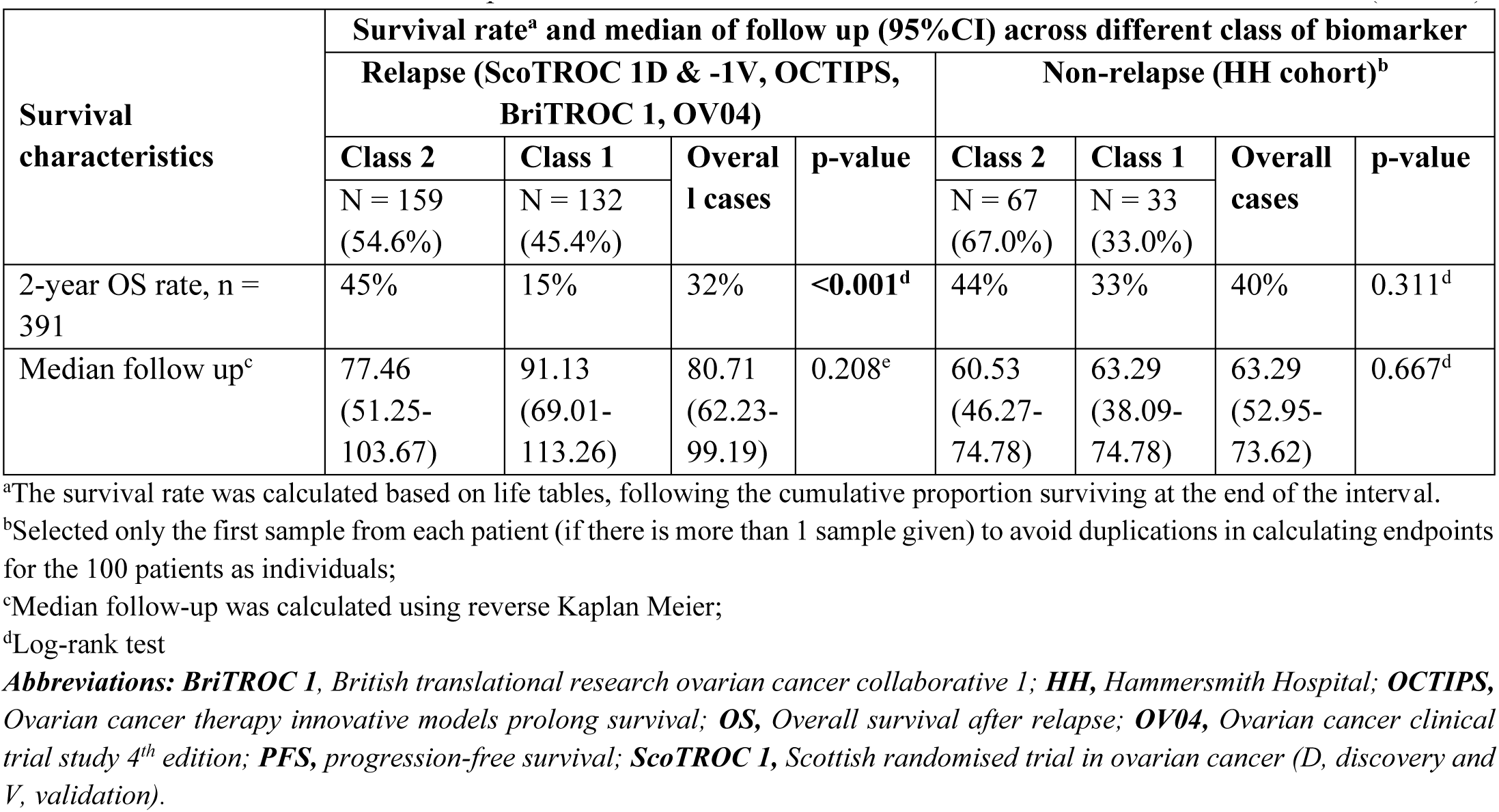
Survival rate differences of patients between two classes of PLAT-M8 in the six included cohorts (n = 391).

**Table S5.** Median progression-free time, time to death, and overall survival time, with a Kaplan-meier survival comparison for relapsed ovarian cancer across various variables.

| Clinicopathological features | Median survival (95%CI), in months |  |  |  |
| --- | --- | --- | --- | --- |
|  | PFS | p-value | OS after relapse | p-value |
| **Biomarker status (at relapse, n = 291) in 5 cohorts** |  |  |  |  |
| Class 2 | 22.59 (19.50-25.67) | <0.001 | 30.44 (22.71-38.17) | <0.001 |
| Class 1 | 11.77 (11.00-12.54) |  | 11.64 (9.58-13.69) |  |
| **Biomarker status (during chemo, n = 100) in HH cohort** |  |  |  |  |
| Class 2 | 23.37 (15.82-30.93) | 0.127 | 28.96 (17.60-40.33) | 0.428 |
| Class 1 | 17.75 (12.45-23.06) |  | 23.01 (11.04-34.99) |  |
| **Age at relapse category (n = 291)** |  |  |  |  |
| Younger (≤75 years) | 16.54 (13.86-19.21) | 0.287 | 20.55 (15.47-25.63) | 0.256 |
| Elder (>75 years) | 18.58 (13.55-23.60) |  | 16.57 (8.90-24.24) |  |
| **FIGO stage degree (n = 291)** |  |  |  |  |
| Early (I-II) | 27.26 (0-55.18) | 0.004 | 46.06 (28.82-63.30) | 0.026 |
| Advance (III-IV) | 15.81 (13.67-17.96) |  | 17.79 (14.17-21.40) |  |
| **Histological group (n = 291)** |  |  |  |  |
| Serous carcinoma | 20.71 (17.23-24.20) | <0.001 | 25.31 (20.28-30.35) | <0.001 |
| Non-serous carcinoma | 11.21 (10.20-12.22) |  | 10.88 (8.33-13.43) |  |
| **First-line chemo (n = 229)** |  |  |  |  |
| Cp monotherapy | 27.81 (0-57.33) | 0.003 | 21.17 (12.07-30.27) | 0.354 |
| Cp +/- other regiments | 24.79 (21.59-27.99) |  | 16.11 (10.50-21.72) |  |
| **Platinum sensitivity (n = 47)** |  |  |  |  |
| Sensitive | 28.54 (22.62-34.45) | <0.001 | 25.74 (16.50-34.98) | <0.001 |
| Resistant | 9.73 (8.89-10.58) |  | 11.14 (2.12-20.17) |  |
| **Surgical type (n = 57)** |  |  |  |  |
| Interval debulking | 22.03 (14.69-29.36) | 0.817 | 21.90 (16.93-26.86) | 0.728 |
| Primary debulking | 20.09 (0.96-39.22) |  | 17.49 (0-44.62) |  |
| **Residual disease (n = 228)** |  |  |  |  |
| No residual disease | 22.03 (17.74-26.31) | <0.001 | 32.61 (24.61-40.62) | <0.001 |
| Any residual disease | 12.00 (11.21-12.79) |  | 13.51 (11.54-15.49) |  |
| **CA-125 response (n = 121)** |  |  |  |  |
| Response (decrease) | 11.93 (11.23-12.64) | 0.004 | 12.62 (9.75-15.49) | 0.697 |
| No response (stable/increase) | 7.60 (1.41-13.678) |  | 8.25 (3.51-12.99) |  |
| **Second-line chemo (n = 104)** |  |  |  |  |
| Cp monotherapy | 26.83 (0-82.35) | 0.064 | 38.00 (16.31-59.70) | 0.238 |
| Other regiments +/- Cp | 22.13 (16.76-27.49) |  | 18.48 (14.14-22.82) |  |
| **Biomarker, second-line chemo (n = 104)** |  |  |  |  |
| Class 2, Cp only | 99.55 (0-205.19) | <0.001 | 38.07 (37.91-38.23) | <0.001 |
| Class 2, others +/- Cp | 24.85 (20.81-28.90) |  | 25.74 (20.44-31.04) |  |
| Class 1, Cp only | 14.79 (8.84-20.75) |  | 11.14 (0.96-12.70) |  |
| Class 1, others +/- Cp | 13.38 (11.35-15.41) |  | 16.57 (13.91-19.22) |  |
| **Biomarker, second-line chemo – Cp only (n = 19)** |  |  |  |  |
| Class 2, Cp only | 99.55 (0-205.19) | 0.001 | 38.07 (37.91-38.23) | <0.001 |
| Class 1, Cp only | 14.79 (8.84-20.75) |  | 11.14 (0.96-12.70) |  |

**Abbreviation:** CA-125, Cancer antigen 125; Cp, carboplatin; FIGO; International federation of gynecology and obstetrics; PFS, Progression-free survival.

**Table S6.**
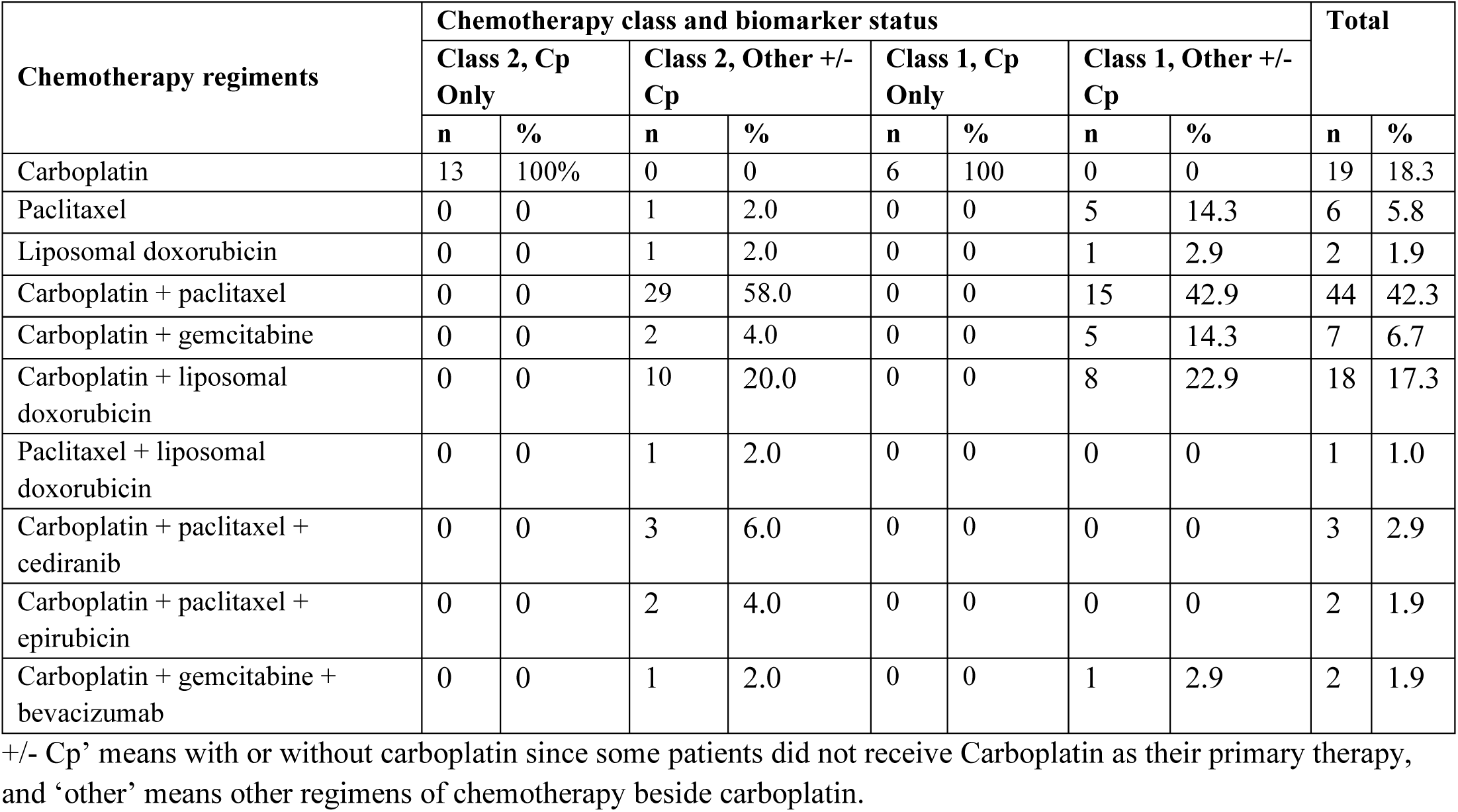
The distribution of second-line chemotherapy regimens based on their grouping by chemotherapy class and biomarker status.

| Chemotherapy regimens | Chemotherapy class and biomarker status |  |  |  |  |  |  |  | Total |  |
| --- | --- | --- | --- | --- | --- | --- | --- | --- | --- | --- |
|  | Class 2, Cp Only |  | Class 2, Other +/- Cp |  | Class 1, Cp Only |  | Class 1, Other +/- Cp |  |  |  |
|  | n | % | n | % | n | % | n | % | n | % |
| Carboplatin | 13 | 100% | 0 | 0 | 6 | 100 | 0 | 0 | 19 | 18.3 |
| Paclitaxel | 0 | 0 | 1 | 2.0 | 0 | 0 | 5 | 14.3 | 6 | 5.8 |
| Liposomal doxorubicin | 0 | 0 | 1 | 2.0 | 0 | 0 | 1 | 2.9 | 2 | 1.9 |
| Carboplatin + paclitaxel | 0 | 0 | 29 | 58.0 | 0 | 0 | 15 | 42.9 | 44 | 42.3 |
| Carboplatin + gemcitabine | 0 | 0 | 2 | 4.0 | 0 | 0 | 5 | 14.3 | 7 | 6.7 |
| Carboplatin + liposomal doxorubicin | 0 | 0 | 10 | 20.0 | 0 | 0 | 8 | 22.9 | 18 | 17.3 |
| Paclitaxel + liposomal doxorubicin | 0 | 0 | 1 | 2.0 | 0 | 0 | 0 | 0 | 1 | 1.0 |
| Carboplatin + paclitaxel + cediranib | 0 | 0 | 3 | 6.0 | 0 | 0 | 0 | 0 | 3 | 2.9 |
| Carboplatin + paclitaxel + epirubicin | 0 | 0 | 2 | 4.0 | 0 | 0 | 0 | 0 | 2 | 1.9 |
| Carboplatin + gemcitabine + bevacizumab | 0 | 0 | 1 | 2.0 | 0 | 0 | 1 | 2.9 | 2 | 1.9 |

+/- Cp' means with or without carboplatin since some patients did not receive Carboplatin as their primary therapy, and 'other' means other regimens of chemotherapy beside carboplatin.

